# Predictors of COVID-19 hospital outcomes: a machine learning analysis of the National COVID Cohort Collaborative

**DOI:** 10.64898/2026.03.06.26347822

**Authors:** Janette Vazquez, Loni Taylor, Yun-Yun K. Chen, Katherine Araya, Madison G. Farnsworth, Xiang Xue, Mahbubul Hasan, N3C Consortium

**Author notes:** Corresponding author: (JV). Membership of the N3C Consortium is provided in the Acknowledgements.

## Abstract

Predicting hospital outcomes for patients with severe acute respiratory infections is critical for risk stratification and resource planning, yet heterogeneous electronic health record (EHR) data, class imbalance, and evolving clinical practice present persistent methodological challenges for machine learning (ML) approaches.

We conducted a retrospective cohort study using EHR data harmonized to the OMOP common data model from the National COVID Cohort Collaborative (N3C; May 2020-June 2025), including 263,619 adults hospitalized with COVID-19 across 51 contributing sites. We developed penalized linear regression (elastic net), random forest, XGBoost, and multilayer perceptron (MLP) models to predict hospital length of stay (LOS) and mortality (in-hospital and 60-day), using demographics, comorbidities, prior healthcare utilization, COVID-19 vaccination status, and hospital site as predictors. Missing data were handled via multiple imputation by chained equations (MICE) and class imbalance was addressed using SMOTE. Model performance was evaluated using area under the ROC curve (AUROC), Brier score, calibration plots, and decision curve analysis, following the TRIPOD reporting framework.

Mortality prediction achieved moderate discrimination across all models (test AUROC = 0.71-0.73 for in-hospital mortality; 0.72-0.73 for 60-day all-cause mortality). Models trained without SMOTE achieved the highest AUROCs but assigned virtually no patients to the mortality class at the default 0.5 threshold. SMOTE improved recall and F-1 score at the cost of reduced AUROC and precision. LOS was poorly explained by available structured predictors (best R^2^ = 0.059). Remdesivir-treated patients (n = 103,536; 39.3%) were older, had higher comorbidity burden, and had higher unadjusted mortality than untreated patients.

Common structured EHR features offer moderate utility for mortality risk stratification in hospitalized COVID-19 patients but are insufficient for LOS prediction. The consistent SMOTE-related tradeoff between discrimination and calibration underscores the need to report threshold-dependent metrics alongside AUROC in clinical ML studies, with implications for operational planning during future respiratory disease emergencies.

## Introduction

Severe acute respiratory infections (SARIs) pose a significant threat to global health, especially during pandemic events when healthcare systems experience heightened strain[1, 2]. The COVID-19 pandemic resulted in hundreds of millions of cases worldwide, overwhelming hospitals and underscoring the urgent need for tools that guide clinical decision-making and support equitable allocation of limited resources[3]. Among the critical determinants of healthcare burden are hospital length of stay (LOS) and patient mortality, two outcomes closely tied to the quality of care, timeliness of clinical interventions, and healthcare costs. Identifying early predictors of these outcomes is vital for informing evidence-based policy and improving patient management during future disease outbreaks (Fig 1).

**Fig 1.**
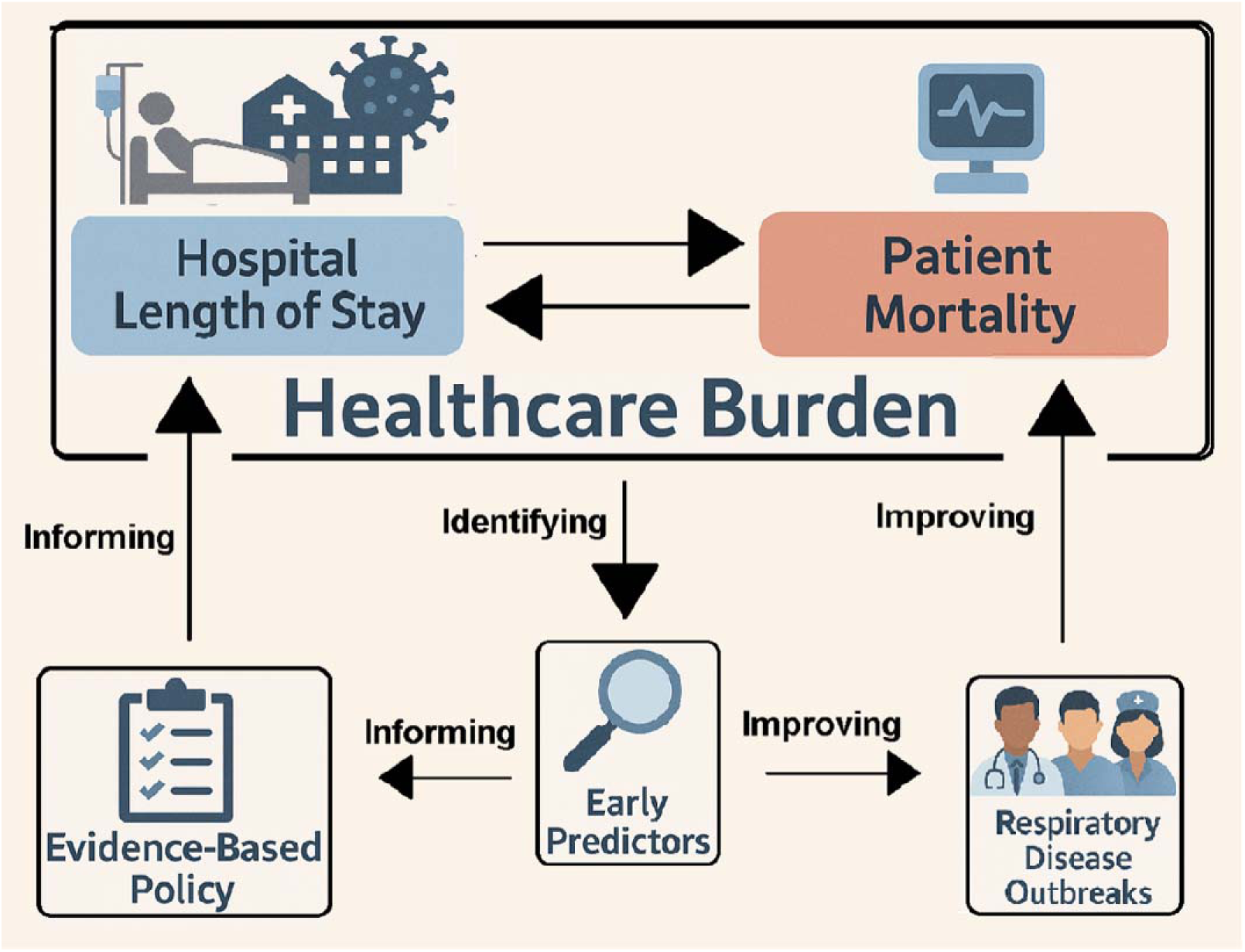
Conceptual framework linking healthcare burden outcomes to evidence-based policy. Hospital length of stay and patient mortality are interrelated drivers of healthcare burden during respiratory disease outbreaks. Identifying early predictors of these outcomes can inform evidence-based policy and improve preparedness for future pandemics.

Antiviral therapies, including remdesivir, have been deployed to mitigate disease severity[4–7]. The ACTT-1 trial demonstrated that remdesivir shortened time to recovery but had no significant effect on mortality in ventilated patients[7]. Systematic reviews indicate that timing of administration, patient characteristics, and comorbidities heavily influence outcomes of antiviral treatments[5, 6, 8]. Remdesivir was the most widely administered inpatient antiviral during the pandemic, yet observational studies report conflicting findings regarding its effectiveness [9, 10], in part because treatment assignment is non-random: clinicians preferentially administered remdesivir to patients perceived as higher risk[11, 12].

Understanding the characteristics of treated versus untreated patients is therefore a necessary first step before applying causal inference methods to estimate treatment effects — and characterizing these differences in large, multi-site cohorts has not been well documented.

Machine learning (ML) approaches have shown promise for clinical outcome prediction using electronic health record (EHR) data[13–15]. However, prior studies have often been limited to single-site cohorts, small sample sizes, or ICU-only populations. The ISARIC 4C Mortality Score, for example, used eight clinical features to achieve an AUROC of 0.77 but was developed using traditional regression[16]. More recent benchmarking efforts have evaluated up to 18 predictive models for COVID-19 outcomes, though primarily in ICU settings with limited generalizability[14].

LOS prediction has proven particularly challenging due to the skewed distribution of stay durations and the influence of unmeasured institutional factors[17, 18]. However, most prior studies have been limited to single-site cohorts or ICU-only populations, and whether these findings generalize across large, heterogeneous EHR datasets remains unclear [17–19]. Whether ML approaches can meaningfully outperform simpler regression models for LOS as a continuous outcome in large, heterogeneous EHR datasets remains an open question.

Despite this growing literature, no study has systematically compared multiple ML architectures for both mortality and LOS prediction using a large, harmonized, multi-site U.S. EHR cohort spanning multiple pandemic waves, while also evaluating the impact of class imbalance correction on both discrimination and calibration. This study aimed to develop and compare ML models for predicting hospital LOS, in-hospital mortality, and 60-day all-cause mortality among adults hospitalized with COVID-19, leveraging the National COVID Cohort Collaborative (N3C), one of the largest harmonized EHR repositories in the United States[20]. We additionally characterize differences in baseline characteristics and unadjusted outcomes by remdesivir exposure to inform the design of future causal analyses. Finally, we evaluate model performance in a pre-specified subgroup of adults aged ≥65 years, a population at disproportionately high risk for severe outcomes[2].

## Methods

We conducted a retrospective cohort study using the N3C EHR repository, which harmonizes data to the Observational Medical Outcomes Partnership (OMOP) common data model[20]. The de identified N3C dataset algorithmically shifts calendar dates per person while preserving within person temporal order.

This study was conducted using de-identified data accessed through the NCATS N3C Data Enclave under a Data Use Agreement between N3C and AIM-AHEAD and an N3C-approved Data Use Request. The N3C data transfer to NCATS is performed under Johns Hopkins University Reliance Protocol # IRB00249128. The project was reviewed and determined not to constitute human subjects research therefore, informed consent was not required. All analyses were performed within the secure N3C Data Enclave.

### Study population

We identified adults ≥18 years admitted with laboratory confirmed SARS CoV 2 infection (positive PCR or standardized N3C diagnosis) within 16 days before hospital admission. We restricted analyses to each patient’s first COVID 19 hospitalization and required LOS > 0 days. We excluded patients who were pregnant, had outpatient only encounters, had missing or inconsistent exposure/outcome timestamps, or were treated at data-partner sites with no inpatient antiviral use during the study window (Fig 2).

**Fig 2.**
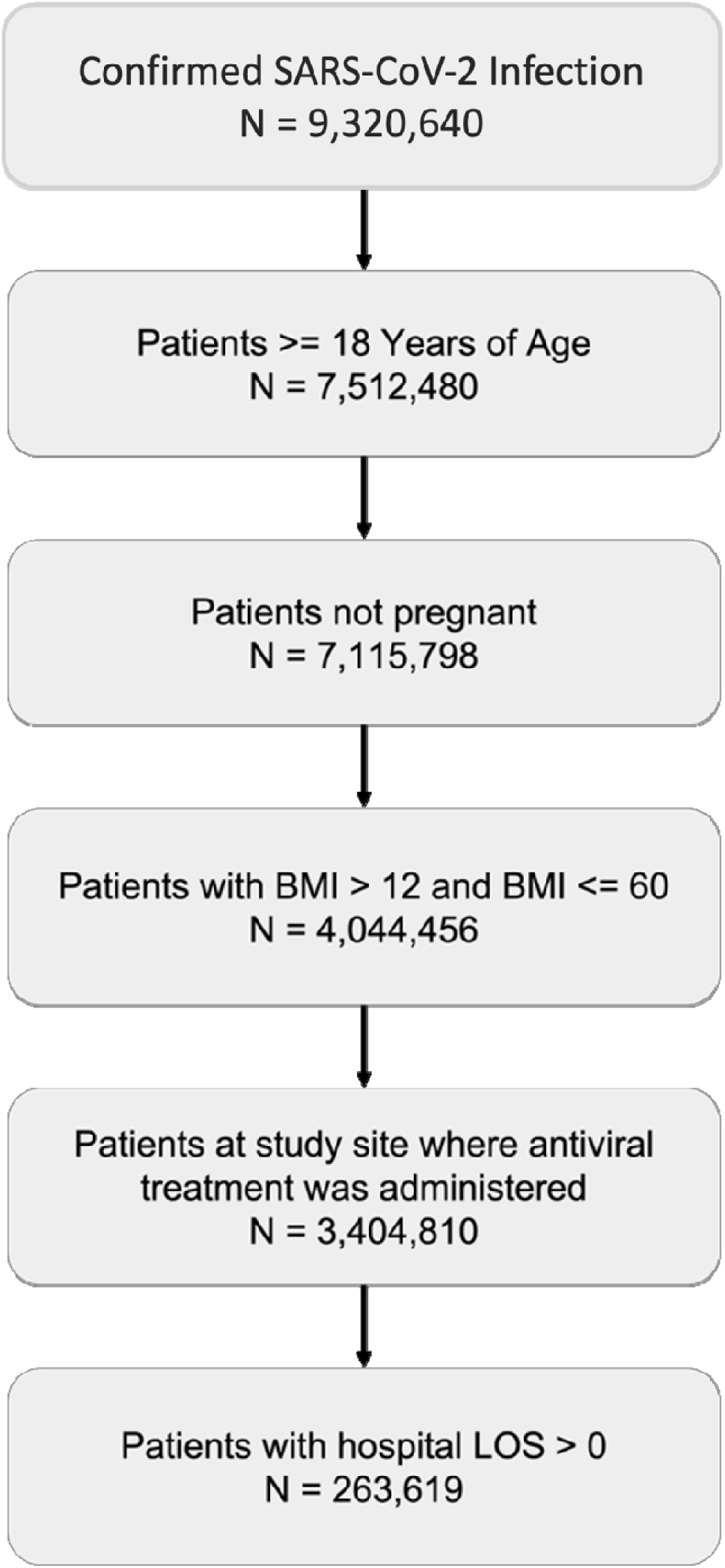
Patient flow diagram. Cohort selection from the National COVID Cohort Collaborative (N3C) Data Enclave, May 2020-June 2025. Beginning with 9,320,640 hospitalized patients with confirmed acute respiratory infection, sequential exclusion criteria were applied: age < 18 years, pregnancy, BMI outside plausible range (<13 or> 60), sites without inpatient antiviral use, and hospital length of stay less than 0 days. The final analytic cohort included 263,619 adults.

### Exposure, outcomes, and predictors

The primary exposure was remdesivir receipt during hospitalization. Outcomes included hospital LOS (continuous, log-transformed for regression), in hospital mortality (death during hospitalization or discharge to hospice), and 60 day all cause mortality (death within 60 days of the admission date). Predictor variables included demographics (age, sex, race/ethnicity), BMI, pre-existing comorbidities recorded on or before admission (hypertension, chronic kidney disease, coronary artery disease, heart failure, cardiomyopathy, cerebrovascular disease, complicated and uncomplicated diabetes, mild and moderate-severe liver disease, malignancy, dementia, depression, substance use, organ transplant, tobacco use, and pulmonary embolism), prior healthcare utilization (counts of visits preceding admission), COVID-19 vaccination status (number of doses), and hospital site identifier.

### Data preprocessing and missing data

Continuous features were standardized (z score) and categorical features were one hot encoded. LOS was log-transformed to reduce skewness. Missing data checks revealed that only number of visits prior to COVID hospitalization had missing values (n=44,504; 16.88%). For this variable, we used Multivariate Imputation by Chained Equations (MICE)[21] to generate a single imputed dataset. Due to platform constraints, we did not pool across multiple imputations, which may understate the uncertainty from missing data. However, the limited extent of missingness (restricted to a single variable) reduces the practical impact of this constraint. We did not conduct a formal missing data mechanism assessment; the MICE procedure assumed data were Missing at Random. Class imbalance for mortality outcomes was addressed using the Synthetic Minority Oversampling Technique (SMOTE)[22] applied within cross validation folds during training.

## Statistical analysis

Baseline characteristics were summarized by treatment group (remdesivir vs no remdesivir) using means with standard deviations for continuous variables and counts with percentages for categorical variables. Group differences were evaluated using t tests and chi-square tests; standardized mean differences (SMDs) were calculated, with an absolute SMD >0.10 considered imbalanced.

### Subgroup analysis

We repeated all predictive modeling analyses in a pre-specified subgroup of adults aged ≥65 years to evaluate whether model performance differs in an older, higher risk population with potentially more homogenous clinical profiles (S1 Table).

### Machine learning models

We developed four model types: (1) penalized linear/logistic regression (elastic net), (2) random forest, (3) XGBoost, and (4) multilayer perceptron (MLP). The dataset was randomly split 80/20 into training and test sets. Hyperparameter tuning was performed via grid search with 3- or 5 fold cross validation on the training data. For classification tasks, models were trained both with and without SMOTE.

Classification performance was evaluated using test area under the Receiver Operating Characteristic curve (AUROC), precision, recall, F1-score, and Brier score. Regression performance was assessed using coefficient of determination (R^2^), root mean square error (RMSE) and mean absolute error (MAE) on the held-out test set. For classification tasks, predicted class labels were assigned using the default probability threshold of 0.5. Threshold optimization was not performed, which may affect sensitivity and specificity estimates, particularly in the presence of class imbalance.

The MLP architecture consisted of a single hidden layer with 50 neurons for classification tasks and 100 neurons for regression, with tanh and relu activation functions, respectively. Hospital site was included as approximately 50 one-hot encoded indicator variables, which constituted a substantial proportion of the total feature space. Feature importance was assessed using permutation importance and SHAP values. All ML models were used for prediction and pattern discovery; we did not interpret their output as causal effects.

Hyperparameters used for our models are reported in the supplementary files (Table S2-S6).

### Software

Data extraction and cohort construction were performed using Spark SQL and PySpark. Statistical analyses and modeling were conducted in R (mice package) and Python (scikit-learn, XGBoost, pandas) within the N3C enclave. The code and analytic workflows are maintained in the enclave for reproducibility.

## Results

### Cohort characteristics

The cohort included 263,619 adults hospitalized with COVID-19 (mean BMI 32.7 kg/m^2^; 47% female) across 51 contributing sites (Table 1). Of these, 103,536 (39.3%) received remdesivir and 160,083 (60.7%) did not. Patients in the remdesivir group were older (mean 63.1 ± 14.7 vs. 59.2 ± 17.1 years; SMD = 0.25), had higher BMI (33.4 ± 8.4 vs. 32.2 ± 8.2; SMD = 0.15), and had more prior healthcare visits (69.2 ± 98.3 vs. 52.5 ± 75.6; SMD = 0.19). Several comorbidities were more prevalent among remdesivir-treated patients, including hypertension (70.2% vs. 62.1%; SMD = 0.17), congestive heart failure (21.5% vs. 16.6%; SMD = 0.12), complicated diabetes (32.7% vs. 27.8%; SMD = 0.11), and coronary artery disease (26.5% vs. 22.1%; SMD = 0.10). Unadjusted mean LOS was similar between groups (9.7 ± 13.4 days vs. 9.8 ± 20.0 days; SMD = 0.01). However, unadjusted in-hospital mortality was higher in the remdesivir group (9.6% vs. 6.6%; SMD = 0.11), as was 60-day mortality (12.5% vs. 9.3%; Table 1).

**Table 1.** Patient characteristics stratified by remdesivir administration - all patients (N=263,619)

| Characteristic | No Remdesivir<br>(N = 160,083) | Remdesivir<br>(N = 103,536) | P-value | SMD |
| --- | --- | --- | --- | --- |
| **Demographics** |  |  |  |  |
| Age (mean ± SD) | 59.2 (±17.1) | 63.1 (±14.7) | <0.001 | 0.249 |
| Sex (female, n, %) | 75,817 (47.4%) | 47,837 (46.2%) | <0.001 | 0.023 |
| BMI (mean ± SD) | 32.2 (±8.2) | 33.4 (±8.4) | <0.001 | 0.151 |
| Obese (n, %) | 93,857 (58.6%) | 67,543 (65.2%) | <0.001 | 0.136 |
| No. Visits before covid<br>(mean ± SD) | 52.5 (±75.6) | 69.2 (±98.3) | <0.001 | 0.191 |
| Race/ethnicity (n, %) | — | — | <0.001 | 0.133 |
| American Indian/Alaska native | 850 (0.5%) | 732 (0.7%) | — | — |
| Asian | 3,958 (2.5%) | 3,056 (3.0%) | — | — |
| Black or African American | 35,557 (22.2%) | 21,860 (21.1%) | — | — |
| Hispanic or Latino (any race) | 17,752 (11.1%) | 10,197 (9.8%) | — | — |
| White non-Hispanic | 93,471 (58.4%) | 62,874 (60.7%) | — | — |
| Other | 2,342 (1.5%) | 3,603 (3.5%) | — | — |
| **Pre-existing conditions (before or day of covid)** |  |  |  |  |
| Hypertension | 99,383 (62.1%) | 72,723 (70.2%) | <0.001 | 0.173 |
| Diabetes (uncomplicated) | 55,576 (34.7%) | 42,093 (40.7%) | <0.001 | 0.123 |
| Diabetes (complicated) | 44,425 (27.8%) | 33,881 (32.7%) | <0.001 | 0.108 |
| Congestive heart failure | 26,590 (16.6%) | 22,223 (21.5%) | <0.001 | 0.124 |
| Coronary artery disease | 35,322 (22.1%) | 27,438 (26.5%) | <0.001 | 0.104 |
| Kidney disease | 41,742 (26.1%) | 30,471 (29.4%) | <0.001 | 0.075 |
| Malignant cancer | 24,114 (15.1%) | 19,211 (18.6%) | <0.001 | 0.093 |
| Cardiomyopathies | 14,394 (9.0%) | 11,481 (11.1%) | <0.001 | 0.070 |
| Cerebrovascular disease | 20,434 (12.8%) | 14,987 (14.5%) | <0.001 | 0.050 |
| Depression | 38,360 (24.0%) | 25,433 (24.6%) | <0.001 | 0.014 |
| Dementia | 10,734 (6.7%) | 7,698 (7.4%) | <0.001 | 0.028 |
| Substance abuse | 16,456 (10.3%) | 7,426 (7.2%) | <0.001 | 0.110 |
| Pulmonary embolism (prior) | 7,485 (4.7%) | 6,794 (6.6%) | <0.001 | 0.082 |
| Myocardial infarction (prior) | 20,727 (12.9%) | 16,379 (15.8%) | <0.001 | 0.082 |
| Solid organ/blood stem cell transplant | 7,379 (4.6%) | 7,206 (7.0%) | <0.001 | 0.101 |
| Tobacco smoker | 28,965 (18.1%) | 20,654 (19.9%) | <0.001 | 0.047 |
| Covid vaccines (n, %) | — | — | <0.001 | 0.049 |
| 0 | 128,306 (80.1%) | 84,499 (81.6%) | — | — |
| 1 | 6,439 (4.0%) | 4,399 (4.2%) | — | — |
| 2+ | 25,338 (15.8%) | 14,638 (14.1%) | — | — |
| **Outcomes** |  |  |  |  |
| Length of stay (mean ± SD, days) | 9.8 (±20.0) | 9.7 (±13.4) | 0.034 | 0.009 |
| In-hospital mortality during covid | 10,597 (6.6%) | 9,970 (9.6%) | <0.001 | 0.110 |
| 60-day all-cause mortality | 14,852 (9.3%) | 12,985 (12.5%) | <0.001 | — |

*Note: SMD = standardized mean difference. Values are mean ± SD or N (%). P-values from chi-square (categorical) or independent samples t-test (continuous). SMD >0.10 generally considered meaningful imbalance.*

In those aged ≥65 years, those treated with remdesivir were generally similar to untreated patients (SMD < 0.10 for most variables), with the exception of prior healthcare visit count (SMD = 0.17), race/ethnicity distribution (SMD = 0.14), and tobacco use (13.6 % smokers in untreated vs 18.9 % smokers in those treated with remdesivir, SMD = 0.14; S1 Table).

### Length of stay prediction

Prediction of log-transformed LOS yielded limited performance across all models in the full cohort. XGBoost achieved the best test-set R^2^ of 0.059 (RMSE = 0.920, MAE = 0.726), followed by random forest (R^2^ = 0.058, RMSE = 0.920) and MLP (R^2^ = 0.055, RMSE = 0.922; Table 2). Elastic net regression performed worst (R^2^ = 0.016, RMSE = 0.941). Cross-validation RMSE values were consistent with test-set results, indicating minimal overfitting. In the ≥65 year subgroup (n = 125, 325), LOS prediction was similarly limited: XGBoost achieved R^2^ = 0.047 (RMSE = 0.872), random forest R^2^ = 0.042, MLP R^2^ = 0.043, and elastic net R^2^ = 0.031 (Table 2). SHAP analyses identified remdesivir treatment, age at COVID-19 diagnosis, complicated diabetes, and prior visit count as the top predictors of LOS in the XGBoost model (Fig 3), with a consistent set of dominant predictors observed across model types (S1–S12 Figs).

**Fig 3.**
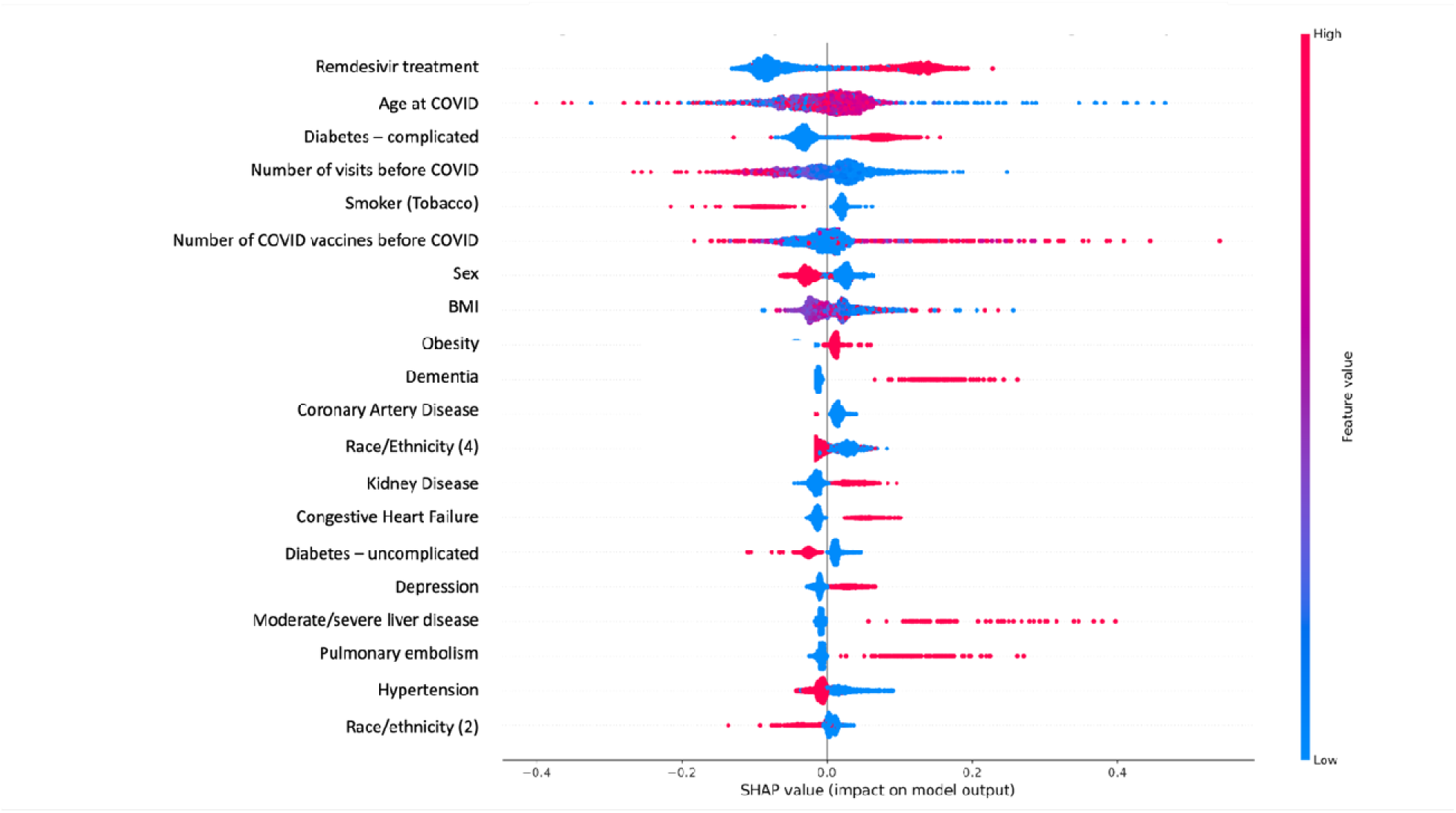
SHAP beeswarm plot for XGBoost regression model predicting log-transformed hospital length of stay (all patients, N = 263,619). Each point represents an individual patient. The horizontal position indicates the SHAP value (impact on model output), and the color indicates the original feature value (red = high, blue = low). Features are ranked by mean absolute SHAP value. Remdesivir treatment, age at COVID-19 diagnosis, complicated diabetes, prior visit count, and tobacco use were the top predictors.

**Table 2.**
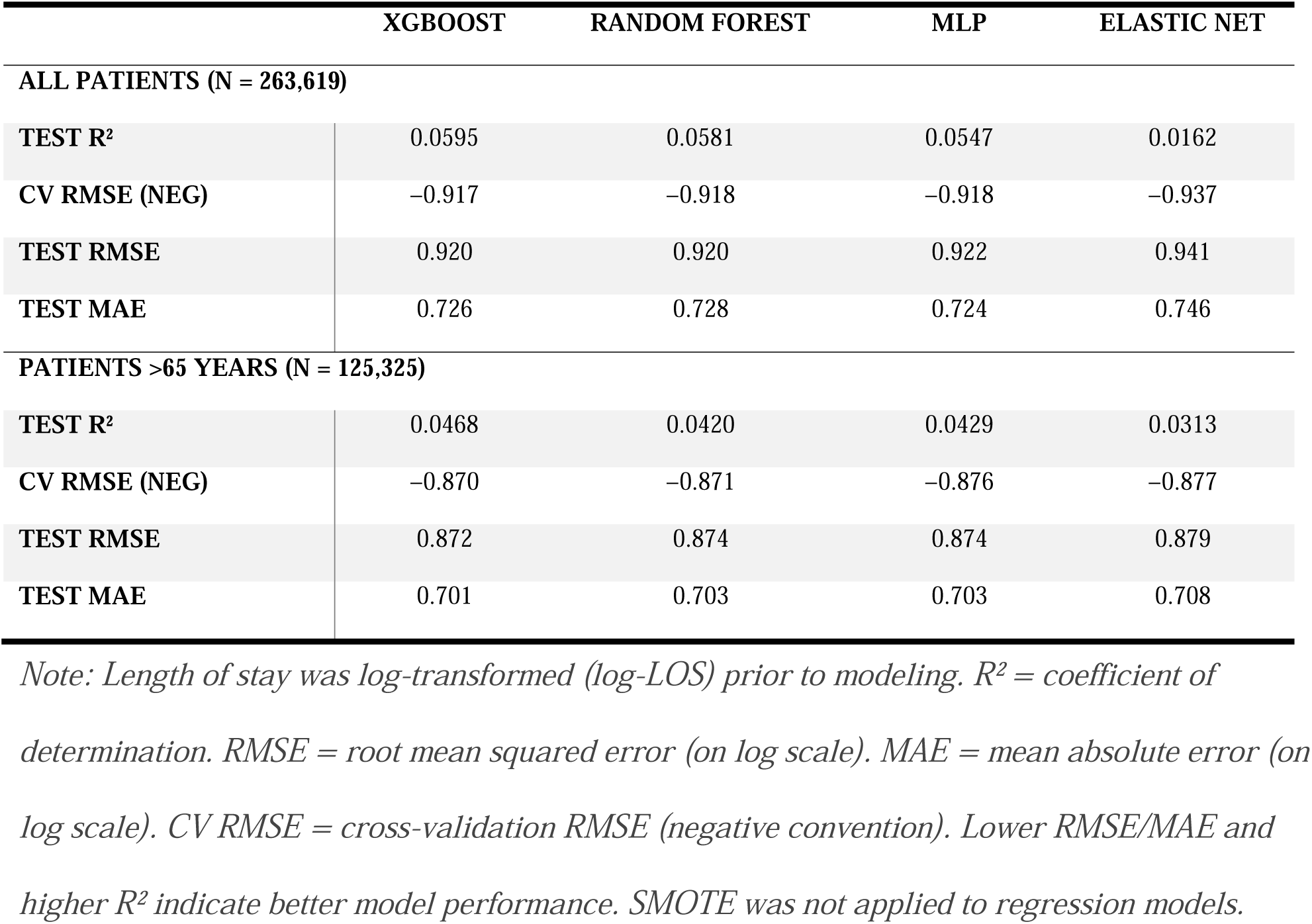
Regression model performance for hospital length of stay - all patients and patients ≥65 years.

|  | XGBOOST | RANDOM FOREST | MLP | ELASTIC NET |
| --- | --- | --- | --- | --- |
| **ALL PATIENTS (N = 263,619)** |  |  |  |  |
| **TEST R<sup>2</sup>** | 0.0595 | 0.0581 | 0.0547 | 0.0162 |
| **CV RMSE (NEG)** | -0.917 | -0.918 | -0.918 | -0.937 |
| **TEST RMSE** | 0.920 | 0.920 | 0.922 | 0.941 |
| **TEST MAE** | 0.726 | 0.728 | 0.724 | 0.746 |
| **PATIENTS >65 YEARS (N = 125,325)** |  |  |  |  |
| **TEST R<sup>2</sup>** | 0.0468 | 0.0420 | 0.0429 | 0.0313 |
| **CV RMSE (NEG)** | -0.870 | -0.871 | -0.876 | -0.877 |
| **TEST RMSE** | 0.872 | 0.874 | 0.874 | 0.879 |
| **TEST MAE** | 0.701 | 0.703 | 0.703 | 0.708 |

*Note: Length of stay was log-transformed (log-LOS) prior to modeling. R<sup>2</sup> = coefficient of determination. RMSE = root mean squared error (on log scale). MAE = mean absolute error (on log scale). CV RMSE = cross-validation RMSE (negative convention). Lower RMSE/MAE and higher R<sup>2</sup> indicate better model performance. SMOTE was not applied to regression models.*

### Mortality prediction

For in-hospital mortality in the full cohort, models trained without SMOTE consistently achieved higher AUROC than SMOTE-augmented models. XGBoost achieved the highest test AUROC without SMOTE (0.721), followed by MLP (0.719; Table 3; Fig 4). Accuracy exceeded 0.92 for all models without SMOTE, reflecting the predominance of survivors. However, F1-scores were near zero and recall was approximately 0, indicating that high AUROC did not translate into the ability to identify individual patients at risk of death. SMOTE improved recall (e.g., random forest recall increased from 0 to 0.59) and F1-score (e.g., random forest F1 increased from 0 to 0.21) at the cost of reduced AUROC and precision (Table 3).

**Fig 4.**
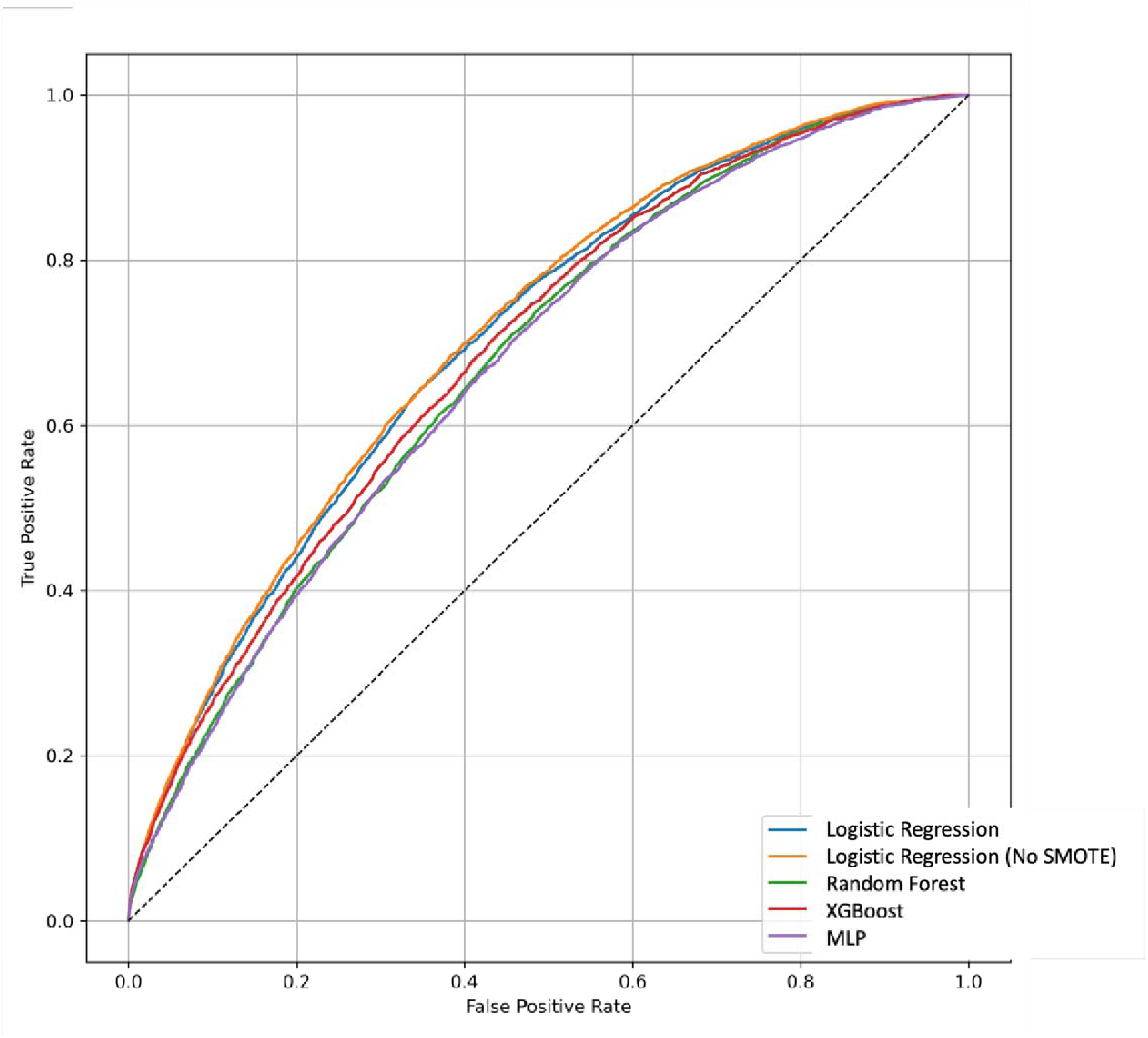
Receiver operating characteristic (ROC) curves for classification models predicting in-hospital mortality (all patients, N = 263,619). Curves are shown for logistic regression with SMOTE (blue), logistic regression without SMOTE (orange), random forest (green), XGBoost (red), and multilayer perceptron (purple). The dashed diagonal represents chance performance (AUROC = 0.50). XGBoost without SMOTE achieved the highest test AUROC of 0.721.

**Table 3.** Classification model performance for mortality outcomes — all patients (N = 263,619)

| Model | SMOTE | CV AUC | Test AUC | F1 | Precision | Recall | BRIER score |
| --- | --- | --- | --- | --- | --- | --- | --- |
| **Outcome: In-hospital mortality** |  |  |  |  |  |  |  |
| **Logistic regression** | No | 0.711 | 0.709 | 0.010 | 0.500 | 0.005 | 0.069 |
|  | Yes | 0.705 | 0.704 | 0.218 | 0.130 | 0.672 | 0.220 |
| **Random forest** | No | 0.715 | 0.713 | 0.000 | — | 0.000 | 0.685 |
|  | Yes | 0.681 | 0.677 | 0.206 | 0.125 | 0.585 | 0.207 |
| **XGBoost** | No | 0.721 | 0.721 | 0.025 | 0.650 | 0.013 | — |
|  | Yes | 0.688 | 0.690 | 0.045 | 0.485 | 0.023 | 0.070 |
| **MLP** | No | 0.719 | 0.719 | 0.021 | 0.597 | 0.010 | — |
|  | Yes | 0.668 | 0.673 | 0.208 | 0.132 | 0.488 | 0.180 |
| **Outcome: 60-day all-cause mortality** |  |  |  |  |  |  |  |
| **Logistic regression** | No | 0.718 | 0.720 | 0.026 | 0.460 | 0.013 | 0.089 |
|  | Yes | 0.712 | 0.715 | 0.281 | 0.177 | 0.679 | 0.218 |
| **Random forest** | No | 0.721 | 0.723 | 0.000 | — | 0.000 | 0.088 |
|  | Yes | 0.692 | 0.695 | 0.273 | 0.185 | 0.514 | 0.180 |
| **XGBoost** | No | 0.727 | 0.731 | 0.041 | 0.646 | 0.021 | — |
|  | Yes | 0.703 | 0.711 | 0.058 | 0.480 | 0.031 | 0.089 |
| **MLP** | No | 0.726 | 0.729 | 0.045 | 0.608 | 0.023 | — |
|  | Yes | 0.682 | 0.695 | 0.271 | 0.209 | 0.387 | 0.154 |

*Note: SMOTE = Synthetic Minority Over-sampling Technique. AUC = Area Under the ROC Curve. Brier Score = mean squared difference between predicted probabilities and outcomes (lower = better). CV AUC = cross-validation AUC on training set. Test AUC = held-out test set AUC. F1 = harmonic mean of precision and recall. '—' indicates value not reported.*

For 60-day all-cause mortality, XGBoost without SMOTE achieved the highest test AUROC (0.731), followed by MLP (0.729), random forest (0.723), and logistic regression (0.720; Table 3; Fig 5). SMOTE improved recall substantially (e.g., logistic regression recall increased from 0.013 to 0.679) but reduced AUROC and precision, mirroring the pattern observed for in-hospital mortality.

**Fig 5.**
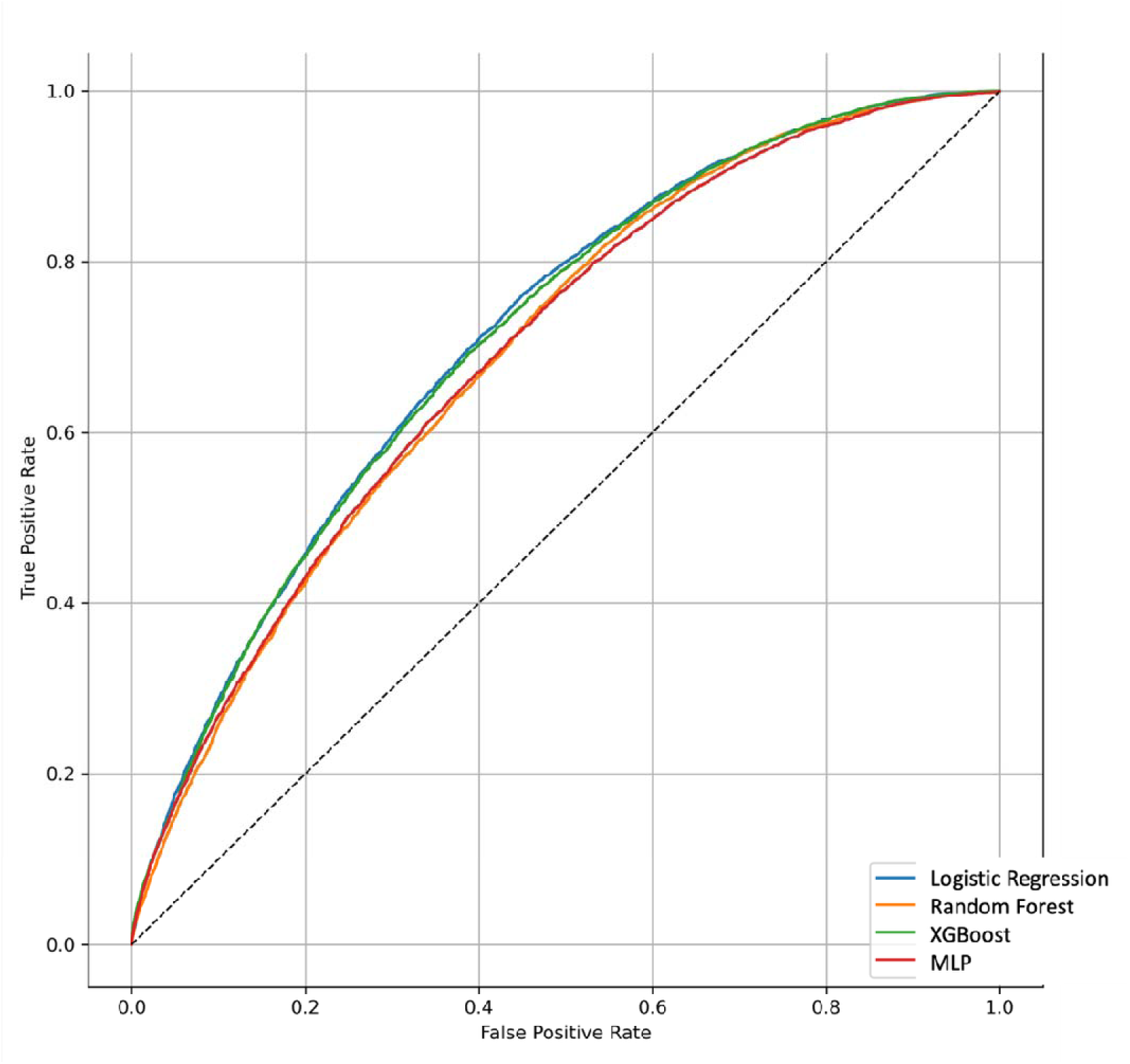
Receiver operating characteristic (ROC) curves for classification models predicting 60-day all-cause mortality (all patients, N = 263, 619). Curves are shown for logistic regression (blue), random forest (orange), XGBoost (green), and multilayer perceptron (red). The dashed diagonal represents chance performance (AUROC = 0.50). XGBoost achieved the highest test AUROC of 0.731.

In the ≥65 year subgroup, mortality discrimination declined meaningfully. For in-hospital mortality, the best models (XGBoost and MLP without SMOTE) achieved an AUROC of 0.661, compared with 0.713 – 0.719 in the full cohort (Table 4). For 60-day all-cause mortality, the best models (XGBoost and MLP without SMOTE) achieved an AUROC of 0.654 (Table 4). LOS prediction remained poor in this subgroup (best R^2^ =0.047; Table 2).

**Table 4.**
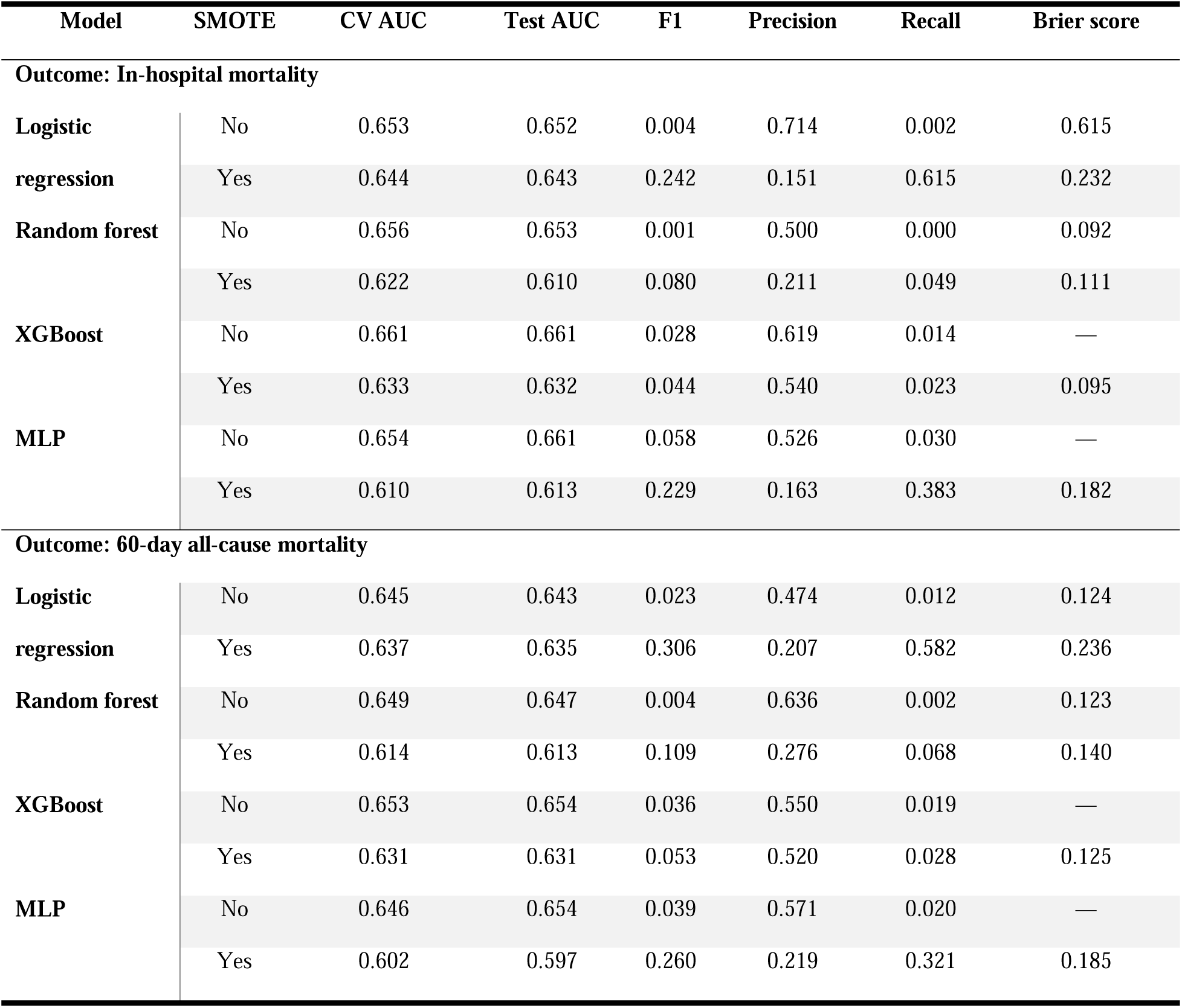
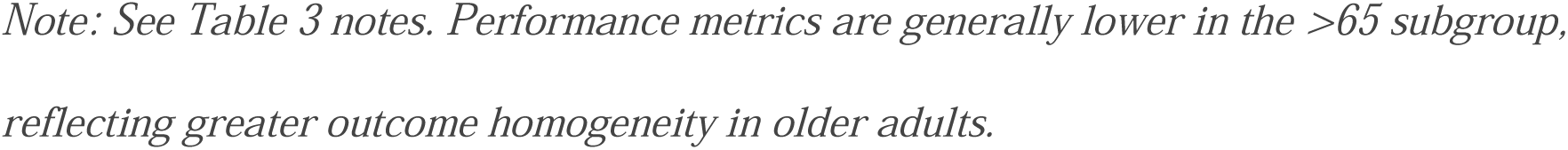
Classification model performance for mortality outcomes - patients >65 Years (N=125,325)

| Model | SMOTE | CV AUC | Test AUC | F1 | Precision | Recall | Brier score |
| --- | --- | --- | --- | --- | --- | --- | --- |
| **Outcome: In-hospital mortality** |  |  |  |  |  |  |  |
| **Logistic regression** | No | 0.653 | 0.652 | 0.004 | 0.714 | 0.002 | 0.615 |
|  | Yes | 0.644 | 0.643 | 0.242 | 0.151 | 0.615 | 0.232 |
| **Random forest** | No | 0.656 | 0.653 | 0.001 | 0.500 | 0.000 | 0.092 |
|  | Yes | 0.622 | 0.610 | 0.080 | 0.211 | 0.049 | 0.111 |
| **XGBoost** | No | 0.661 | 0.661 | 0.028 | 0.619 | 0.014 | — |
|  | Yes | 0.633 | 0.632 | 0.044 | 0.540 | 0.023 | 0.095 |
| **MLP** | No | 0.654 | 0.661 | 0.058 | 0.526 | 0.030 | — |
|  | Yes | 0.610 | 0.613 | 0.229 | 0.163 | 0.383 | 0.182 |
| **Outcome: 60-day all-cause mortality** |  |  |  |  |  |  |  |
| **Logistic regression** | No | 0.645 | 0.643 | 0.023 | 0.474 | 0.012 | 0.124 |
|  | Yes | 0.637 | 0.635 | 0.306 | 0.207 | 0.582 | 0.236 |
| **Random forest** | No | 0.649 | 0.647 | 0.004 | 0.636 | 0.002 | 0.123 |
|  | Yes | 0.614 | 0.613 | 0.109 | 0.276 | 0.068 | 0.140 |
| **XGBoost** | No | 0.653 | 0.654 | 0.036 | 0.550 | 0.019 | — |
|  | Yes | 0.631 | 0.631 | 0.053 | 0.520 | 0.028 | 0.125 |
| **MLP** | No | 0.646 | 0.654 | 0.039 | 0.571 | 0.020 | — |
|  | Yes | 0.602 | 0.597 | 0.260 | 0.219 | 0.321 | 0.185 |

*Note: See Table 3 notes. Performance metrics are generally lower in the >65 subgroup, reflecting greater outcome homogeneity in older adults.*

### Model calibration

Calibration plots for SMOTE-augmented mortality models revealed moderate miscalibration across all model types (S1–S8 Figs). Models generally tracked the calibration diagonal at lower predicted probabilities but overestimated mortality risk at moderate-to-high predicted values (approximately 0.3–0.6 predicted probability range). Among SMOTE-augmented models, the 60-day mortality calibration curves more closely followed the diagonal than those for in-hospital mortality. Decision curve analyses indicated that model-based risk stratification provided positive net benefit over default strategies (treat-all or treat-none) at threshold probabilities below approximately 0.2, corresponding to the clinically relevant range for screening applications.

## Discussion

In this large, multi-site study of 263,619 adults hospitalized with COVID-19, we found that ML models achieved moderate discrimination for mortality prediction (AUROC 0.71-0.73; Table 3) but poor performance for LOS prediction (R^2^ < 0.06; Table 2) using structured EHR data. These results are consistent with prior work demonstrating that LOS is inherently difficult to predict using baseline patient-level features alone[17, 18]. Hospital level factors such as institutional discharge protocols, bed capacity, staffing ratios, and regional practice variation likely account for substantial LOS variability that is not captured by patient-level predictors. The identification of hospital site as a top predictor of LOS reinforces this interpretation and underscores the importance of including facility-level variables in predictive models.

Our mortality AUROCs of 0.71-0.73 are somewhat lower than the 0.77 reported for the ISARIC 4C Mortality Score[16], though direct comparison is difficult given differences in cohort composition, time period, and predictor sets. The 4C score incorporated physiological measures at admission (e.g., respiratory rate, oxygen saturation, Glasgow Coma Scale) that were not available as structured variables in our analytic dataset. The absence of real-time clinical severity markers, viral strain information, and symptom onset timing likely constrained model performance. These findings align with benchmarking studies that found moderate discrimination to be typical for EHR-based COVID-19 prediction tasks, particularly when restricted to structured data[14, 23].

Beyond discrimination, we evaluated model calibration using calibration plots and decision curve analyses (S1–S8 Figs). SMOTE-augmented models exhibited moderate miscalibration, particularly overestimating risk at moderate-to-high predicted probabilities. This pattern is consistent with the known tendency of SMOTE to inflate predicted probabilities by altering the class distribution in the training data[22]. Models trained without SMOTE were well-calibrated within their narrow prediction range but were clinically limited in their ability to identify high-risk patients, as predicted probabilities rarely exceeded the observed event rate.

These calibration findings underscore that discrimination (AUROC) alone is insufficient for evaluating clinical prediction models - a model may rank patients correctly while systematically overestimating or underestimating absolute risk, which has direct implications for clinical decision thresholds and resource allocation[24].

A notable finding was the reduced model performance in adults aged ≥65 years, where the best AUROC for 60-day all-cause mortality declined from 0.73 in all patients to 0.65 in the age-restricted subgroup (Table 3; Table 4). This likely reflects greater homogeneity in risk profiles among older adults; when most patients share similarly elevated comorbidity burdens and advanced age, the discriminative power of these same features diminishes. This has practical implications for risk stratification in elderly populations, where complementary data sources (e.g., frailty indices, functional status, laboratory trajectories) may be needed to achieve clinically useful prediction.

Remdesivir-treated patients were older, had more comorbidities, and experienced higher unadjusted mortality than untreated patients (Table 1), differences consistent with confounding by indication[11, 12]. These imbalances (age SMD = 0.25, BMI SMD = 0.15, hypertension SMD = 0.17, and prior healthcare utilization SMD = 0.19) characterize the selection process underlying treatment assignment and preclude causal conclusions from naive group comparisons. We present this descriptive stratification to document the nature and magnitude of confounding in a large, multi-site U.S. cohort, informing the design of future causal studies using propensity score methods, target trial emulation, or instrumental variable designs [11, 25, 26].

The SMOTE analyses revealed an important methodological tradeoff, oversampling improved recall and F1-scores for the minority class but consistently reduced AUROC and precision. Notably, several models trained without SMOTE using the default 0.5 classification threshold effectively never predicted death as the positive class, yielding recall values of zero. This dissociation between discrimination and classification carries broader implications for the reporting of ML models in clinical research. A model that achieves a respectable AUROC, indicating correct relative ranking of patient risk, but classifies no patients as high-risk at the default decision threshold provides no actionable clinical information at point of care. This pattern was consistent across all four model architectures for both mortality outcomes, suggesting it is a property of outcome imbalance rather than a model-specific artifact. These results underscore that AUROC alone is an insufficient metric for evaluating prediction models applied to imbalanced clinical outcomes[22] and that threshold-dependent metrics (precision, recall, F1-score) and calibration assessments should be reported alongside discrimination in all clinical ML studies. The optimal modeling strategy ultimately depends on the intended use case: population-level risk ranking (e.g., triage dashboards) may favor models maximizing AUROC regardless of threshold behavior, whereas bedside screening applications require reliable identification of high-risk individuals, potentially through SMOTE augmentation, threshold optimization, cost-sensitive learning, or alternative loss functions.

Ensemble ML methods (XGBoost, random forest) generally outperformed logistic regression, though margins were modest (XGBoost AUROC of 0.731 vs. logistic regression AUROC of 0.720 for 60-day mortality; Table 3), consistent with systematic reviews showing that ML methods often provide only incremental improvements over well-specified regression models when using similar feature sets[23, 24]. The MLP models used relatively simple architectures (single hidden layer, 50 neurons) and may benefit from deeper networks in future analyses, though at the cost of greater computational demands and overfitting risk. The greatest advantage of tree-based models may lie in their ability to capture nonlinear interactions and produce interpretable feature importance rankings rather than in raw discriminative performance. In our analysis, these models consistently identified age, hospital site, and comorbidity burden as dominant predictors of both LOS and mortality, findings that are clinically plausible and consistent with prior COVID-19 literature[14, 16, 27]. Notably, feature importance rankings were consistent across model types, with age, hospital site, and comorbidity burden (complicated diabetes, moderate-to-severe liver disease, and kidney disease) as dominant predictors of both LOS and mortality regardless of model type. This convergence suggests that the identified predictors reflect genuine signal in the data rather than model-specific artifacts, and aligns with established evidence that metabolic and organ dysfunction comorbidities are key determinants of COVID-19 severity[14, 27].

The identification of hospital site as a top predictor of both LOS and mortality warrants further consideration. Hospital site was included as approximately 50 one-hot encoded indicator variables, which constituted a substantial proportion of the total feature space. Tree-based models (XGBoost, random forest) can preferentially select among these many site-level indicators at each split, which may partly inflate the apparent importance of hospital site relative to individual clinical predictors. Nevertheless, the finding is clinically meaningful: site-level variation likely captures institutional discharge protocols, bed capacity, staffing ratios, and regional practice patterns that genuinely drive outcome heterogeneity beyond patient-level factors. Future analyses should consider mixed-effects or hierarchical modeling frameworks that explicitly separate within-site and between-site variation, which may provide more transportable predictions and more accurate estimates of patient-level predictor importance.

Our study has several strengths, including the large sample size, multi-site design spanning five years of data across 51 U.S. institutions, use of harmonized EHR data through the OMOP common data model, and systematic comparison of multiple ML approaches with and without class imbalance correction. By including a pre-specified age-stratified subgroup analysis, we provide evidence on the generalizability of ML models across clinically relevant populations. We reported model performance following the Transparent Reporting of a multivariable prediction model for Individual Prognosis Or Diagnosis (TRIPOD) framework where possible, including discrimination, calibration, and decision curve analyses. Regarding practical implications, these findings suggest that mortality risk scores derived from structured EHR features could be integrated into hospital dashboards for early identification of high-risk patients, though the modest discrimination highlights the need for additional data inputs (e.g., laboratory trajectories, imaging findings, genomic data) to achieve clinically actionable performance[24, 28]. Any deployment should be accompanied by prospective validation, calibration assessment, and impact evaluation.

### Limitations

This study has several limitations. First, the retrospective design introduces the possibility of residual confounding and selection bias. Critical variables, including symptom onset timing, viral strain, detailed admission severity scores (e.g., oxygen saturation, respiratory rate), and prehospital treatments, were unavailable in the analytic dataset. Second, we used a single imputed dataset rather than pooling multiple imputations due to computational constraints, which may underestimate uncertainty.

Third, we did not perform temporal validation or pandemic wave-stratified analyses. The study period (May 2020 - June 2025) spans the transition from wild-type SARS-CoV-2 through Alpha, Delta, and multiple Omicron sub-lineages, alongside major shifts in vaccination coverage, treatment protocols, and population immunity. Model performance, predictor-outcome relationships, and the underlying mortality base rate likely varied substantially across these periods, and the pooled estimates may mask such temporal heterogeneity. The N3C date-shifting algorithm, which shifts calendar dates per patient to protect de-identification while preserving within-person temporal order, prevents exact alignment of patients to specific calendar periods or pandemic waves, limiting the feasibility of time-stratified analyses. Relatedly, vaccination status was treated as a static baseline variable in our models, but it is inherently entangled with calendar time. Patients hospitalized before vaccine availability (2020) necessarily had zero doses, whereas unvaccinated patients hospitalized in 2022 or later represent a distinct subpopulation that may differ systematically in health-seeking behavior, comorbidity burden, and access to care. This time-varying confounding may influence both the apparent predictive value of vaccination status and its interaction with other predictors. Future work should evaluate temporal model stability, ideally using external datasets in which calendar time is available.

Fourth, we did not perform external validation outside the N3C consortium, which limits generalizability. Fifth, hospital site was included as approximately 50 one-hot encoded variables. This high-dimensional representation may overstate the relative importance of site-level effects in tree-based models and limits the transportability of predictions to new institutions. While SHAP values and feature importance were computed, we did not conduct subgroup-level fairness analyses or evaluate model performance across racial/ethnic groups. Given that the cohort included approximately 22% Black or African American and 10-11% Hispanic patients, subgroup-specific model performance (AUROC and calibration by race/ethnicity) would be informative for assessing whether these models perform equitably across populations, a critical consideration before any clinical deployment [24]. Future work should evaluate algorithmic fairness metrics and examine whether predictor importance varies across racial/ethnic subgroups. Finally, these ML models identify associations and improve prediction but do not establish causation. Prospective studies or formal causal inference methods would be needed to confirm treatment effects [25, 26].

## Conclusion

In this large, multi-site, EHR-derived cohort of 263,619 hospitalized COVID-19 patients, mortality outcomes were moderately predictable using common structured features (AUROC 0.71–0.73), whereas LOS remained poorly explained by baseline patient-level and treatment variables (R² < 0.06). SHAP-based feature importance analyses identified age, BMI, prior healthcare utilization, complicated diabetes, moderate-to-severe liver disease, and kidney disease as consistent predictors across model types, supporting the clinical plausibility of these findings. Calibration analyses revealed that models trained with SMOTE tended to overestimate risk at moderate-to-high predicted probabilities, while models trained without SMOTE failed to classify any patients as high-risk at the default probability threshold despite achieving the highest AUROCs. Together, these findings reinforce that discrimination metrics alone are insufficient for evaluating clinical prediction models with imbalanced outcomes, and that threshold-dependent metrics and calibration assessments are essential reporting elements.

Remdesivir exposure was associated with higher unadjusted mortality consistent with confounding by indication, and the baseline differences documented here can inform the design of future causal inference studies. Model performance was notably reduced among adults aged ≥65 years, underscoring the need for age-specific approaches and richer predictor sets with functional status, laboratory trajectories, and frailty measures for risk stratification in elderly populations. These findings illustrate both the potential and the recognized limitations of structured EHR data for predictive modeling in pandemic settings and suggest that meaningful improvements in prediction will require integration of clinical severity markers, temporal disease dynamics, and facility-level factors not captured in routine structured data.

## Data Availability

The data underlying this study are available through the National COVID Cohort Collaborative (N3C) Data Enclave, which is maintained by the National Center for Advancing Translational Sciences (NCATS). Access to N3C data requires registration and approval through the N3C Data Access Committee. Requests for access can be submitted at https://ncats.nih.gov/n3c/about/applying-for-access. The analysis code used in this study is available upon reasonable request to the corresponding author.

## Acknowledgements

The analyses described in this publication were conducted with data or tools accessed through the NCATS N3C Data Enclave (https://covid.cd2h.org) and N3C Attribution & Publication Policy v1.2-2020-08-25b supported by NCATS U24 TR002306. This research was possible because of the patients whose information is included within the data and the organizations (https://ncats.nih.gov/n3c/resources/data-contribution/data-transfer-agreement-signatories) and scientists who have contributed to the on-going development of this community resource.

## Disclaimer

The content is solely the responsibility of the authors and does not necessarily represent the official views of the National Institutes of Health or the N3C program.

We gratefully acknowledge the following core contributors to N3C: Adam B. Wilcox, Adam M. Lee, Alexis Graves, Alfred (Jerrod) Anzalone, Amin Manna, Amit Saha, Amy Olex, Andrea Zhou, Andrew E. Williams, Andrew Southerland, Andrew T. Girvin, Anita Walden, Anjali A. Sharathkumar, Benjamin Amor, Benjamin Bates, Brian Hendricks, Brijesh Patel, Caleb Alexander, Carolyn Bramante, Cavin Ward-Caviness, Charisse Madlock-Brown, Christine Suver, Christopher Chute, Christopher Dillon, Chunlei Wu, Clare Schmitt, Cliff Takemoto, Dan Housman, Davera Gabriel, David A. Eichmann, Diego Mazzotti, Don Brown, Eilis Boudreau, Elaine Hill, Elizabeth Zampino, Emily Carlson Marti, Emily R. Pfaff, Evan French, Farrukh M Koraishy, Federico Mariona, Fred Prior, George Sokos, Greg Martin, Harold Lehmann, Heidi Spratt, Hemalkumar Mehta, Hongfang Liu, Hythem Sidky, J.W. Awori Hayanga, Jami Pincavitch, Jaylyn Clark, Jeremy Richard Harper, Jessica Islam, Jin Ge, Joel Gagnier, Joel H. Saltz, Joel Saltz, Johanna Loomba, John Buse, Jomol Mathew, Joni L. Rutter, Julie A. McMurry, Justin Guinney, Justin Starren, Karen Crowley, Katie Rebecca Bradwell, Kellie M. Walters, Ken Wilkins, Kenneth R. Gersing, Kenrick Dwain Cato, Kimberly Murray, Kristin Kostka, Lavance Northington, Lee Allan Pyles, Leonie Misquitta, Lesley Cottrell, Lili Portilla, Mariam Deacy, Mark M. Bissell, Marshall Clark, Mary Emmett, Mary Morrison Saltz, Matvey B. Palchuk, Melissa A. Haendel, Meredith Adams, Meredith Temple-O’Connor, Michael G. Kurilla, Michele Morris, Nabeel Qureshi, Nasia Safdar, Nicole Garbarini, Noha Sharafeldin, Ofer Sadan, Patricia A. Francis, Penny Wung Burgoon, Peter Robinson, Philip R.O. Payne, Rafael Fuentes, Randeep Jawa, Rebecca Erwin-Cohen, Rena Patel, Richard A. Moffitt, Richard L. Zhu, Rishi Kamaleswaran, Robert Hurley, Robert T. Miller, Saiju Pyarajan, Sam G. Michael, Samuel Bozzette, Sandeep Mallipattu, Satyanarayana Vedula, Scott Chapman, Shawn T. O’Neil, Soko Setoguchi, Stephanie S. Hong, Steve Johnson, Tellen D. Bennett, Tiffany Callahan, Umit Topaloglu, Usman Sheikh, Valery Gordon, Vignesh Subbian, Warren A. Kibbe, Wenndy Hernandez, Will Beasley, Will Cooper, William Hillegass, Xiaohan Tanner Zhang. Details of contributions available at covid.cd2h.org/core-contributors.

This work was supported by the AIM-AHEAD Coordinating Center, funded by the NIH. The authors gratefully acknowledge the Axle Informatics Data Science Education Team, including Aubri Hoffman, Alen Delic, Jide Sadiq, and Hadis Hashemi, for their scholarly contributions and engagement during the Fireside Session and the Annual In-Person Meeting.

We also extend our appreciation to the Axle Instructional Design (ID) team for their coordination and professional support in facilitating these academic activities.

AI-assisted editing tools (ChatGPT, OpenAI, accessed [02/2026]) were used to improve grammar and clarity of selected passages. All content was verified and approved by the authors, who take full responsibility for the accuracy of the manuscript.

## Supporting information

**S1 Table.** Patient characteristics stratified by remdesivir administration – patients aged ≥65 years (N = 125, 325).

**S2 Table.** Best hyperparameters for logistic regression models predicting in-hospital mortality and 60-day all-cause mortality.

**S3 Table.** Best hyperparameters for Random Forest models predicting in-hospital mortality and 60-day all-cause mortality.

**S4 Table**. Best hyperparameters for XGBoost models predicting in-hospital mortality and 60-day all-cause mortality.

**S5 Table.** Multilayer perceptron architecture and training parameters for classification tasks predicting in-hospital mortality and 60-day all-cause mortality.

**S6 Table.** Best hyperparameters for regression models predicting length of stay.

**S1 Fig.** Random Forest model calibration plot, decision curve, and mean SHAP feature importance for in-hospital mortality with and without SMOTE (all patients).

**S2 Fig.** XGBoost model calibration plot, decision curve, and mean SHAP feature importance for in-hospital mortality with SMOTE (all patients).

**S3 Fig.** Multilayer perceptron model calibration plot, decision curve, training loss curve, and mean SHAP feature importance for in-hospital mortality with SMOTE (all patients).

**S4 Fig.** Logistic regression model calibration plot, decision curve, and SHAP beeswarm plot for in-hospital mortality with SMOTE (all patients).

**S5 Fig.** Random Forest model calibration plot, decision curve, and mean SHAP feature importance for 60-day mortality with SMOTE (all patients).

**S6 Fig.** XGBoost model calibration plot, decision curve, and mean SHAP feature importance for 60-day mortality with SMOTE (all patients).

**S7 Fig.** Multilayer perceptron model calibration plot, decision curve, training loss curve, and mean SHAP feature importance for 60-day mortality with SMOTE (all patients).

**S8 Fig.** Logistic regression model calibration plot, decision curve, and SHAP beeswarm plot for 60-day mortality with SMOTE (all patients).

**S9 Fig.** Elastic nets model calibration plot, diagnostic plot, and mean SHAP feature importance for length of stay (all patients).

**S10 Fig.** Random Forest model calibration plot, diagnostic plot, and mean SHAP feature importance for length of stay (all patients).

**S11 Fig.** XGBoost model calibration plot and diagnostic plot for length of stay (all patients).

**S12 Fig.** Multilayer perceptron model plots for length of stay (all patients).

**S13 Fig.** AUROC comparison of classification models for in-hospital mortality (all patients). **S14 Fig.** AUROC comparison of classification models for 60-day all-cause mortality on all patients.

