## SupplementalFile for "Predictors of COVID-19 hospital outcomes: a machine learning analysis of the National COVID Cohort Collaborative"

**SUPPLEMENTARY MATERIAL**

### S1 Table. Patient Characteristics Stratified by Remdesivir Administration — Patients ≥65 Years (N = 125,325)

| Characteristic | No Remdesivir  (N = 71,047) | Remdesivir  (N = 54,278) | P-Value | | SMD |
| --- | --- | --- | --- | --- | --- |
| Demographics | | | | | |
| Age (Mean ± SD) | 74.35 (±5.84) | 74.42 (±5.88) | 0.031 | 0.012 | |
| Sex (FEMALE, N, %) | 35,029 (49.3%) | 25,903 (47.7%) | <0.001 | 0.034 | |
| BMI (Mean ± SD) | 31.45 (±7.49) | 32.05 (±7.64) | <0.001 | 0.079 | |
| Obese (N, %) | 40,090 (56.4%) | 32,378 (59.7%) | <0.001 | 0.065 | |
| No. Visits Before COVID (Mean ± SD) | 61.23 (±81.0) | 77.31 (±102.1) | <0.001 | 0.174 | |
| Length of Stay — log (Mean ± SD) | 1.67 (±0.92) | 1.88 (±0.83) | <0.001 | 0.236 | |
| Length of Stay — days (Mean ± SD) | 8.49 (±12.95) | 9.59 (±11.76) | <0.001 | 0.089 | |
| Race/Ethnicity (N, %) | — | — | <0.001 | 0.139 | |
| American Indian/Alaska Native | 291 (0.4%) | 271 (0.5%) | — | — | |
| Asian Non-Hispanic | 1,875 (2.6%) | 1,705 (3.1%) | — | — | |
| Black or African American | 12,874 (18.1%) | 9,054 (16.7%) | — | — | |
| Hispanic or Latino (Any race) | 5,426 (7.6%) | 3,621 (6.7%) | — | — | |
| Native Hawaiian/Pacific Islander | 320 (0.5%) | 280 (0.5%) | — | — | |
| Other Non-Hispanic | 1,031 (1.5%) | 173 (0.3%) | — | — | |
| Unknown | 2,135 (3.0%) | 1,653 (3.0%) | — | — | |
| White Non-Hispanic | 47,095 (66.3%) | 37,521 (69.1%) | — | — | |
| No. COVID Vaccines (N, %) | — | — | <0.001 | 0.043 | |
| 0 | 54,007 (76.0%) | 41,185 (75.9%) | — | — | |
| 1 | 3,039 (4.3%) | 2,681 (4.9%) | — | — | |
| 2 | 6,046 (8.5%) | 4,787 (8.8%) | — | — | |
| 3 | 4,562 (6.4%) | 3,328 (6.1%) | — | — | |
| 4 | 3,393 (4.8%) | 2,297 (4.2%) | — | — | |
| Pre-existing Conditions (Before or Day of COVID) | | | | | |
| Hypertension | 55,983 (78.8%) | 44,298 (81.6%) | <0.001 | 0.071 | |
| Diabetes (Uncomplicated) | 30,173 (42.5%) | 24,447 (45.0%) | <0.001 | 0.052 | |
| Diabetes (Complicated) | 24,332 (34.2%) | 19,971 (36.8%) | <0.001 | 0.053 | |
| Congestive Heart Failure | 17,058 (24.0%) | 15,301 (28.2%) | <0.001 | 0.095 | |
| Coronary Artery Disease | 24,474 (34.4%) | 20,235 (37.3%) | <0.001 | 0.059 | |
| Kidney Disease | 25,691 (36.2%) | 20,245 (37.3%) | <0.001 | 0.024 | |
| Malignant Cancer | 15,574 (21.9%) | 13,357 (24.6%) | <0.001 | 0.064 | |
| Metastatic Solid Tumor Cancer | 199 (0.3%) | 265 (0.5%) | <0.001 | 0.034 | |
| Cardiomyopathies | 8,326 (11.7%) | 7,227 (13.3%) | <0.001 | 0.048 | |
| Cerebrovascular Disease | 14,198 (20.0%) | 11,207 (20.6%) | 0.004 | 0.016 | |
| Depression | 16,183 (22.8%) | 14,100 (26.0%) | <0.001 | 0.075 | |
| Dementia | 9,629 (13.6%) | 7,034 (13.0%) | 0.002 | 0.018 | |
| Mild Liver Disease | 7,675 (10.8%) | 6,386 (11.8%) | <0.001 | 0.030 | |
| Moderate to Severe Liver Disease | 2,271 (3.2%) | 1,798 (3.3%) | 0.257 | 0.007 | |
| Substance Abuse | 3,341 (4.7%) | 2,539 (4.7%) | 0.848 | 0.001 | |
| Pulmonary Embolism (prior) | 3,641 (5.1%) | 3,773 (7.0%) | <0.001 | 0.077 | |
| Myocardial Infarction (prior) | 12,777 (18.0%) | 11,141 (20.5%) | <0.001 | 0.065 | |
| Solid Organ/Blood Stem Cell Transplant | 3,361 (4.7%) | 3,646 (6.7%) | <0.001 | 0.086 | |
| Tobacco Smoker | 9,677 (13.6%) | 10,234 (18.9%) | <0.001 | 0.142 | |
| Outcomes | | | | | |
| Length of Stay — days (Mean ± SD) | 8.49 (±12.95) | 9.59 (±11.76) | <0.001 | 0.089 | |
| Death During COVID Hospitalization | 6,868 (9.7%) | 6,589 (12.1%) | <0.001 | 0.079 | |
| Death Within Specified Window | 10,023 (14.1%) | 8,919 (16.4%) | <0.001 | 0.065 | |

*Note: SMD = standardized mean difference. Values are mean ± SD or N (%). P-values from chi-square (categorical) or independent samples t-test (continuous). SMD >0.10 generally considered meaningful imbalance.*

### S2 Table. Best hyperparameters for logistic regression models predicting in-hospital mortality and 60-day all-cause mortality.

| **Hyperparameter** | **All Patients** | | | | **Patients >65** | | | |
| --- | --- | --- | --- | --- | --- | --- | --- | --- |
|  | **Death: COVID No SMOTE** | **Death: COVID SMOTE** | **Death: Window No SMOTE** | **Death: Window SMOTE** | **Death: COVID No SMOTE** | **Death: COVID SMOTE** | **Death: Window No SMOTE** | **Death: Window SMOTE** |
| Penalty | L1 | L1 | L1 | L2 | L2 | L2 | L2 | L2 |
| C (Regularization) | 0.167 | 0.167 | 100.0 | 27.83 | 0.013 | 0.013 | 0.599 | 0.013 |

*Note: Penalty = regularization type (L1=Lasso, L2=Ridge). C = inverse of regularization strength (higher = less regularization). Tuned via 3-fold cross-validation with 10-candidate random search. Death:COVID = in-hospital mortality; Death: Window = 60-day all-cause mortality.*

### S3 Table. Best hyperparameters for Random Forest models predicting in-hospital and 60-day all-cause mortality.

| Hyperparameter | All Patients | | | | Patients >65 | | | |
| --- | --- | --- | --- | --- | --- | --- | --- | --- |
|  | **Death: COVID No SMOTE** | **Death: COVID SMOTE** | **Death: Window No SMOTE** | **Death: Window SMOTE** | **Death: COVID No SMOTE** | **Death: COVID SMOTE** | **Death: Window No SMOTE** | **Death: Window SMOTE** |
| n_estimators | 200 | 200 | 200 | 200 | 200 | 100 | 200 | 100 |
| min_samples_split | 5 | 2 | 2 | 5 | 5 | 10 | 2 | 10 |
| min_samples_leaf | 2 | 4 | 4 | 1 | 2 | 2 | 4 | 2 |
| max_features | log2 | sqrt | log2 | log2 | log2 | sqrt | log2 | sqrt |
| max_depth | 10 | 5 | 10 | 10 | 10 | None | 10 | None |

*Note: max_depth=None indicates trees grown until all leaves are pure. Tuned via 3-fold cross-validation with 10-candidate random search. Death:COVID = in-hospital mortality; Death: Window = 60-day all-cause mortality.*

### S4 Table. Best hyperparameters for XGBoost models predicting in-hospital mortality and 60-day all-cause mortality.

| Hyperparameter | All Patients | | | | Patients >65 | | | |
| --- | --- | --- | --- | --- | --- | --- | --- | --- |
|  | **Death: COVID No SMOTE** | **Death: COVID SMOTE** | **Death: Window No SMOTE** | **Death: Window SMOTE** | **Death: COVID No SMOTE** | **Death: COVID SMOTE** | **Death: Window No SMOTE** | **Death: Window SMOTE** |
| n_estimators | 600 | 600 | 600 | 600 | 600 | 400 | 600 | 400 |
| max_depth | 7 | 5 | 7 | 5 | 7 | 5 | 7 | 5 |
| learning_rate | 0.01 | 0.20 | 0.01 | 0.20 | 0.01 | 0.10 | 0.01 | 0.10 |
| subsample | 0.8 | 0.8 | 0.8 | 0.8 | 0.8 | 0.8 | 0.8 | 0.8 |
| min_child_weight | 3 | 1 | 3 | 1 | 3 | 5 | 3 | 5 |
| colsample_bytree | 0.6 | 0.6 | 0.6 | 0.6 | 0.6 | 0.8 | 0.6 | 0.8 |

*Note: learning_rate = step size shrinkage. subsample = fraction of samples per tree. colsample_bytree = fraction of features per tree. min_child_weight = minimum sum of instance weight needed in a child. Tuned via 3-fold cross-validation with 10-candidate random search. Death:COVID = in-hospital mortality; Death: Window = 60-day all-cause mortality.*

### S5 Table. Multilayer perceptron (MLP) architecture and training parameters for classification tasks predicting in-hospital mortality and 60-day all-cause mortality.

| Hyperparameter | All Patients | | | | Patients >65 | | | |
| --- | --- | --- | --- | --- | --- | --- | --- | --- |
|  | **Death: COVID No SMOTE** | **Death: COVID SMOTE** | **Death: Window No SMOTE** | **Death: Window SMOTE** | **Death: COVID No SMOTE** | **Death: COVID SMOTE** | **Death: Window No SMOTE** | **Death: Window SMOTE** |
| solver | adam | adam | adam | adam | adam | adam | adam | adam |
| learning_rate_init | 0.001 | 0.010 | 0.001 | 0.010 | 0.010 | 0.010 | 0.001 | 0.010 |
| hidden_layer_sizes | (50,) | (50,) | (50,) | (50,) | (50,) | (50,) | (50,) | (50,) |
| alpha | 0.01 | 0.01 | 0.01 | 0.01 | 0.01 | 0.01 | 0.01 | 0.01 |
| activation | tanh | tanh | tanh | tanh | tanh | tanh | tanh | tanh |

*Note: solver=adam in all conditions. alpha = L2 regularization term. hidden_layer_sizes=(50,) = single hidden layer with 50 neurons. Tuned via 3-fold cross-validation with 10-candidate random search. Death:COVID = in-hospital mortality; Death: Window = 60-day all-cause mortality.*

### S6 Table. Best hyperparameters for regression models predicting length of stay.

| Hyperparameter | Random Forest | XGBoost | MLP | Elastic Net |
| --- | --- | --- | --- | --- |
| All Patients (N = 263,619) | | | | |
| n_estimators | 300 | 500 | — | — |
| max_depth | 20 | 4 | — | — |
| min_samples_split | 2 | — | — | — |
| min_samples_leaf | 4 | — | — | — |
| max_features | sqrt | — | — | — |
| learning_rate | — | 0.05 | 0.005† | — |
| subsample | — | 0.7 | — | — |
| min_child_weight | — | 5 | — | — |
| colsample_bytree | — | 1.0 | — | — |
| gamma | — | 0.1 | — | — |
| hidden_layer_sizes | — | — | (100,) | — |
| alpha | — | — | 0.01 | 0.00774 |
| activation | — | — | relu | — |
| l1_ratio | — | — | — | 0.75 |
| Patients >65 Years (N = 125,325) | | | | |
| *Identical hyperparameters to All Patients cohort (tuning yielded same optimal values)* | | | | |

*Note: LOS models. Elastic Net alpha = overall regularization strength; l1_ratio = mixing parameter (1.0 = Lasso, 0.0 = Ridge). '—' = hyperparameter not applicable to that model. Patients >65 tuning yielded identical optimal hyperparameters to the full cohort.
† MLP learning_rate_init (initial learning rate).*

### S1 Fig. Random Forest model (a) calibration plot, (b) decision curve, and (c) mean SHAP feature importance for in-hospital mortality during COVID-19 hospitalization with SMOTE and without SMOTE (all patients).

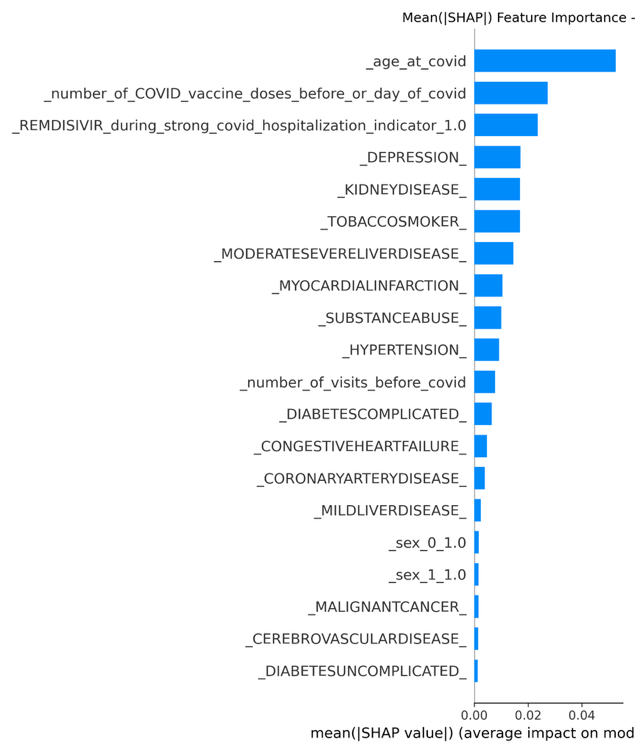

**With SMOTE:**

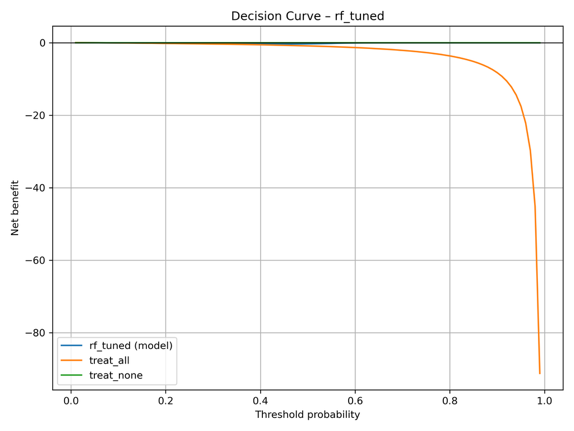

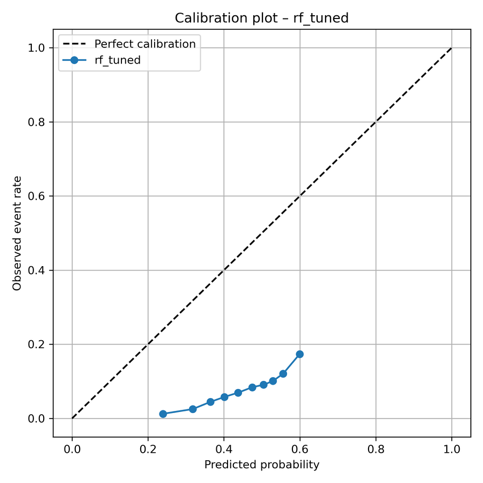

**( b )**

**( a )**

**( c )**

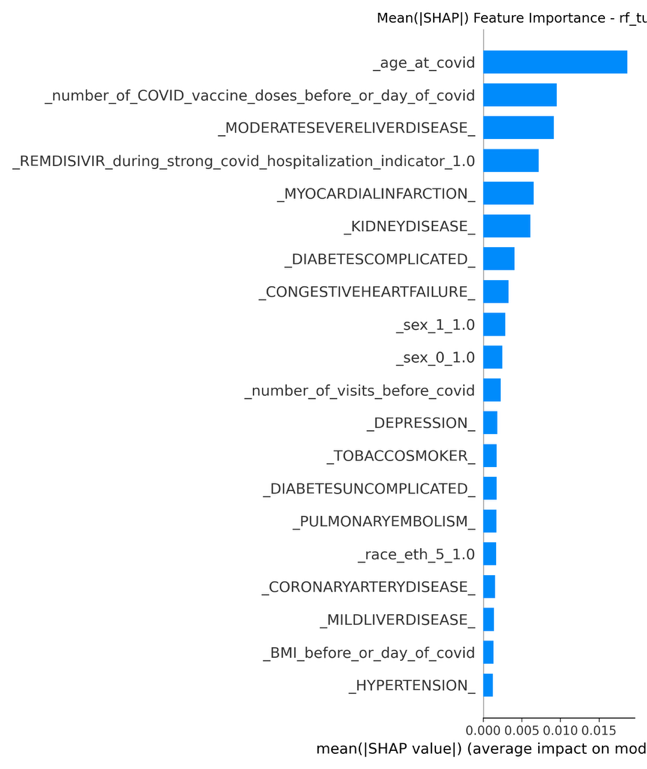

**Without SMOTE:**

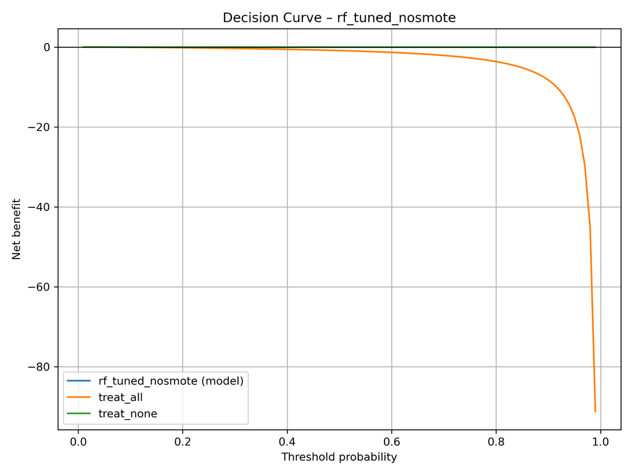

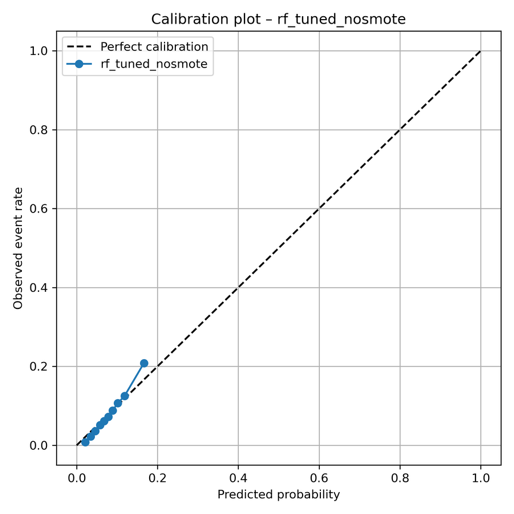

**( b )**

**( a )**

**( c )**

### S2 Fig. XGBoost model (a) calibration plot, (b) decision curve, and (c) mean SHAP feature importance for in-hospital mortality during COVID-19 hospitalization with SMOTE (all patients).

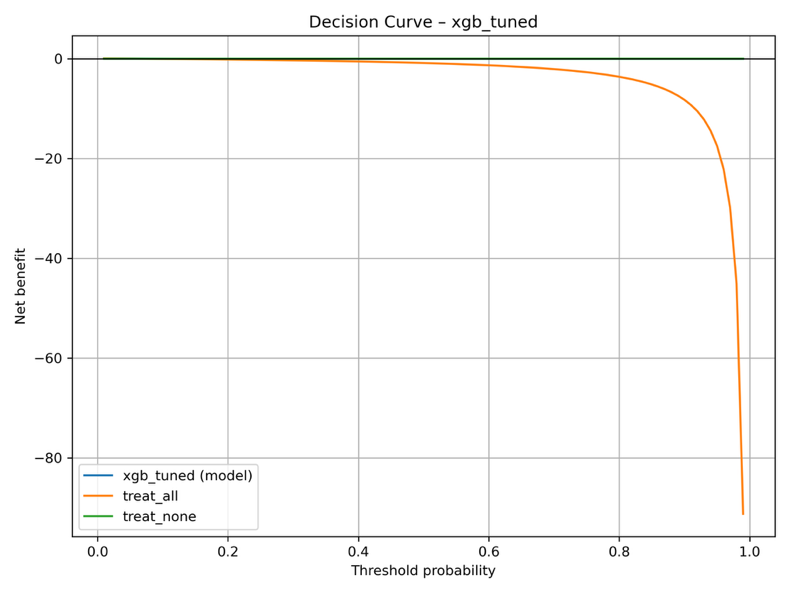

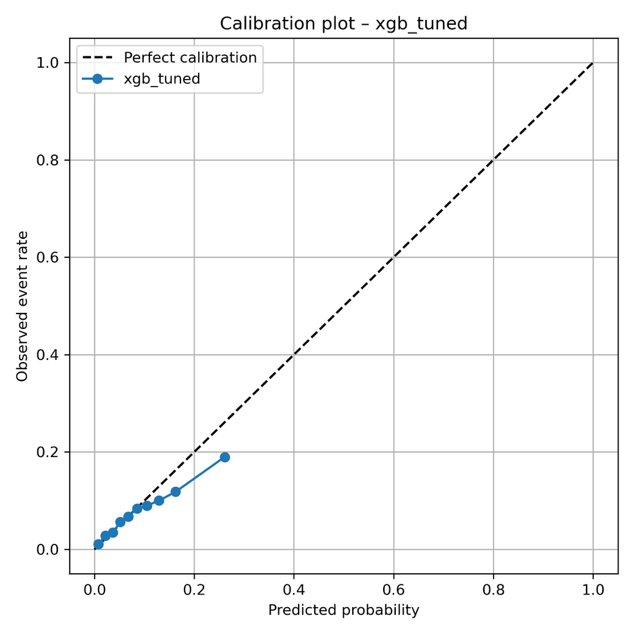

**( b )**

**( a )**

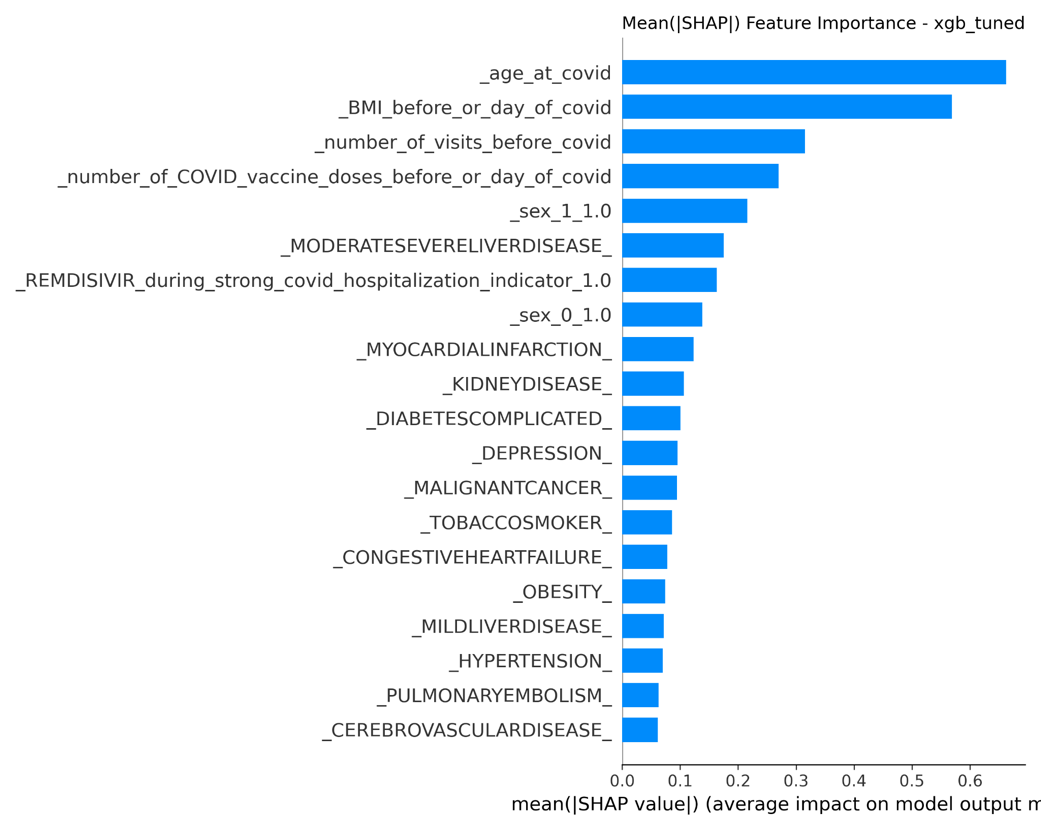

**( c )**

### S3 Fig. Multilayer perceptron (MLP) model (a) calibration plot, (b) decision curve, (c) training loss curve, and (d) mean SHAP feature importance for in-hospital mortality during COVID-19 hospitalization with SMOTE (all patients).

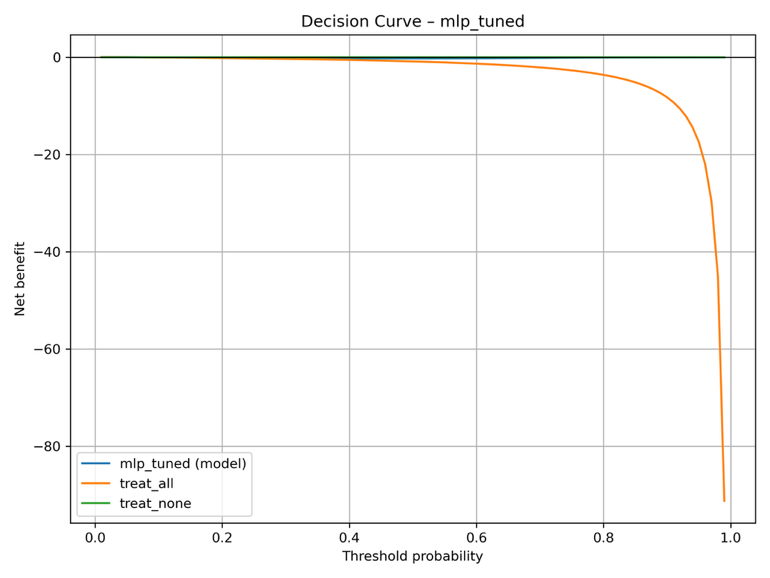

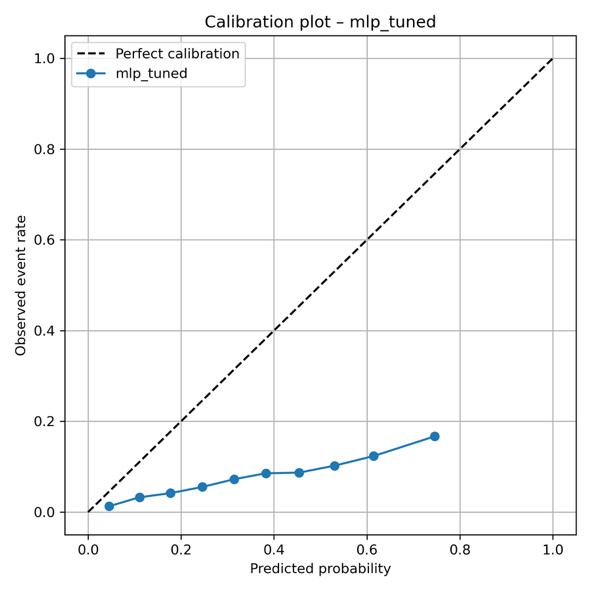

**( b )**

**( a )**

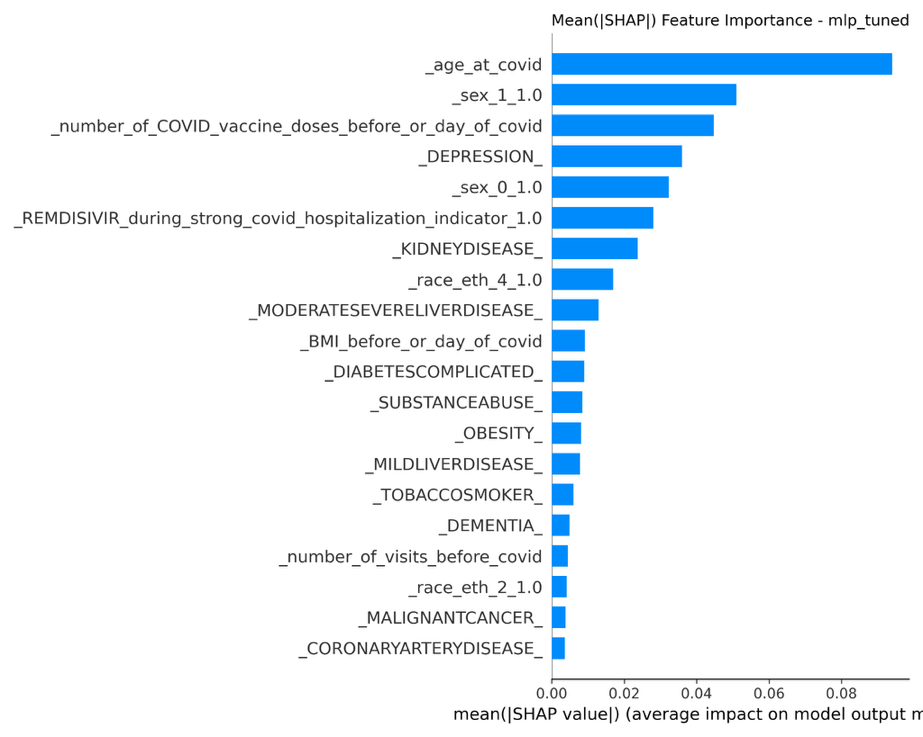

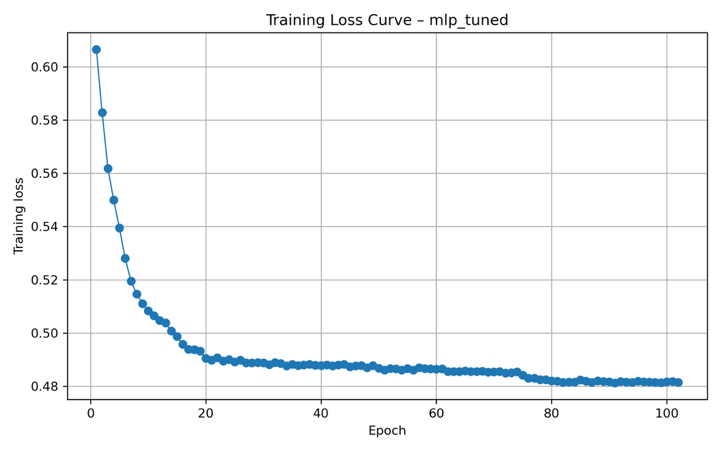

**( c )**

**( d )**

### S4 Fig. Logistic regression model (a) calibration plot, (b) decision curve, and (c) SHAP beeswarm plot for in-hospital mortality during COVID-19 hospitalization with SMOTE (all patients).

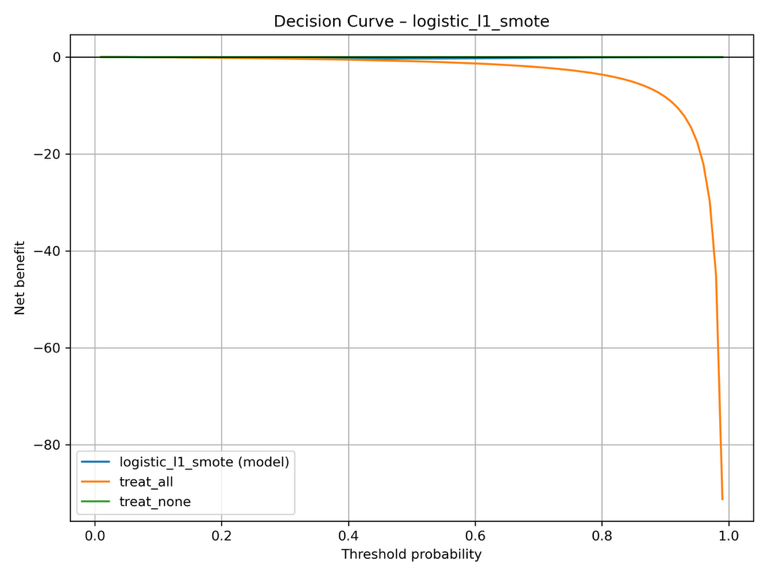

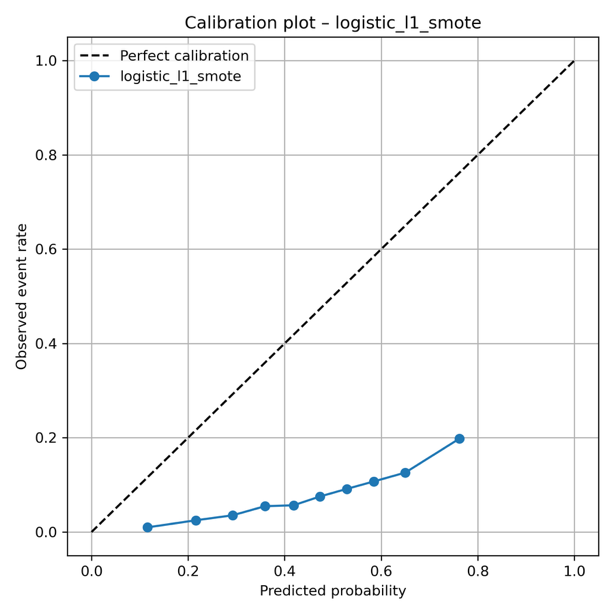

**( b )**

**( a )**

**( c )**

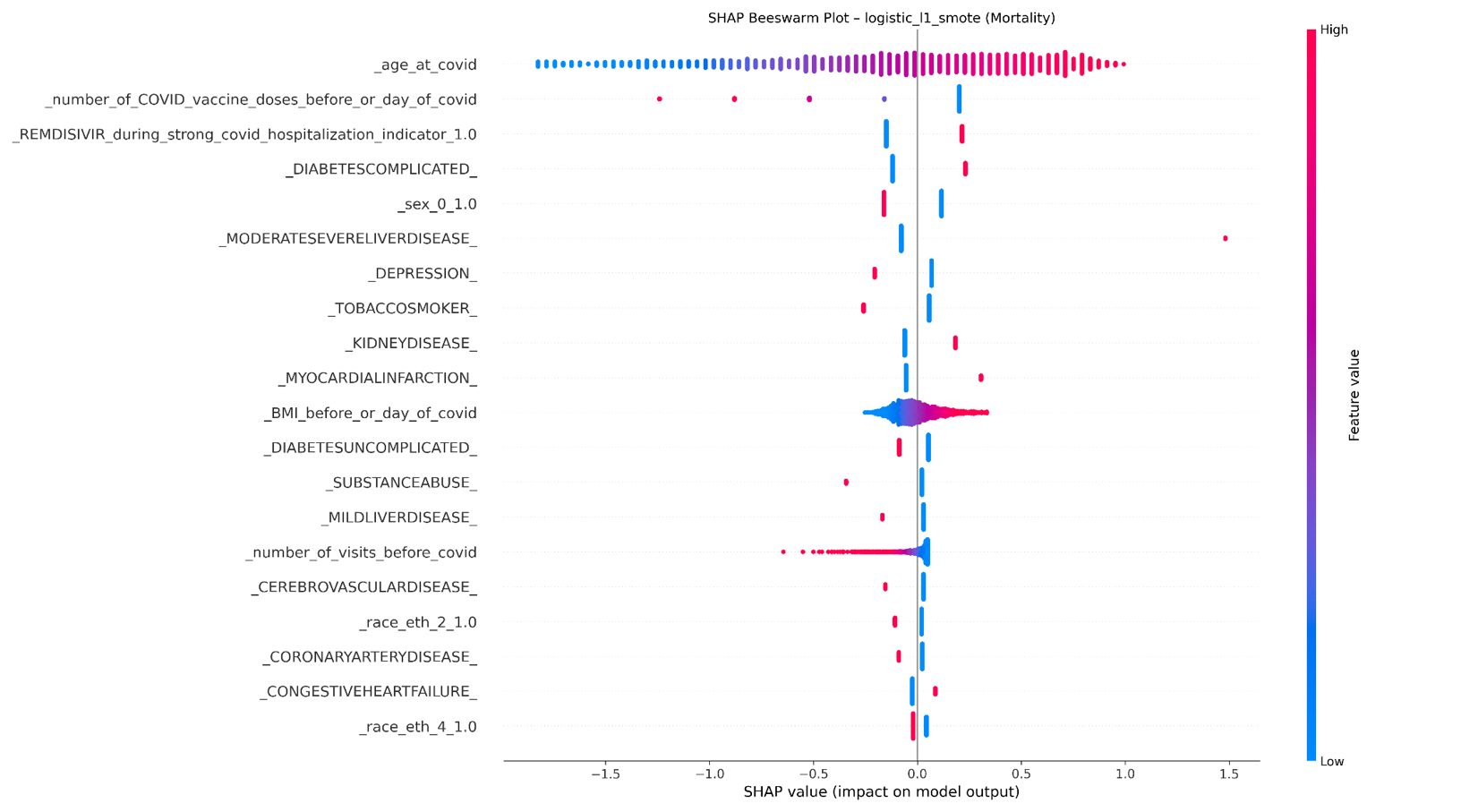

### S5 Fig. Random Forest model (a) calibration plot, (b) decision curve, and (c) mean SHAP feature importance for 60-day mortality with SMOTE (all patients).

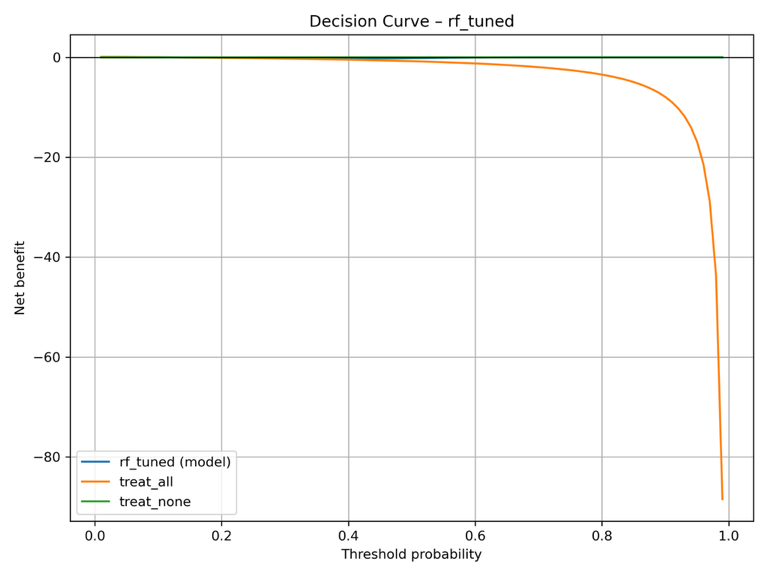

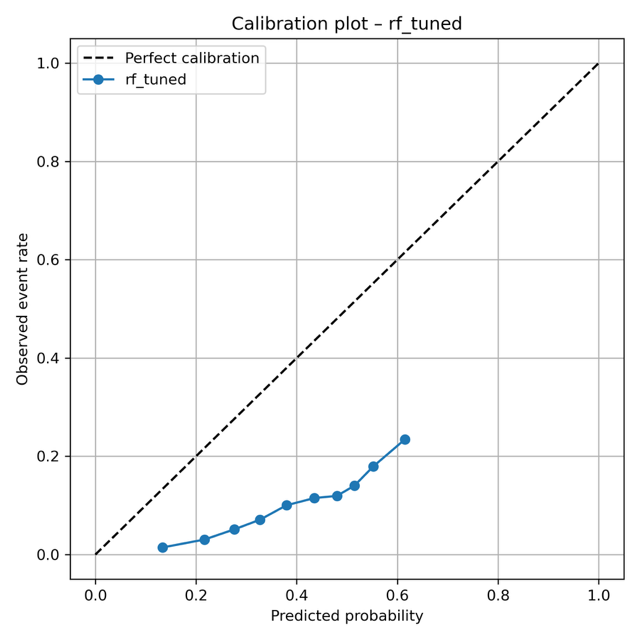

**( a )**

**( b )**

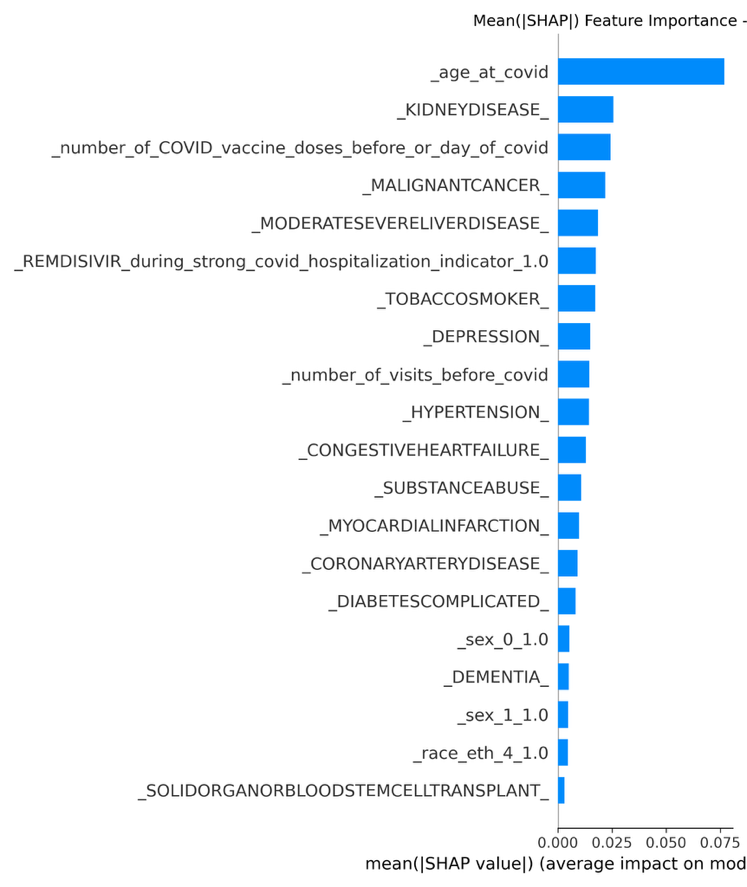

**( c )**

### S6 Fig. XGBoost model (a) calibration plot, (b) decision curve, and (c) mean SHAP feature importance for 60-day mortality with SMOTE (all patients).

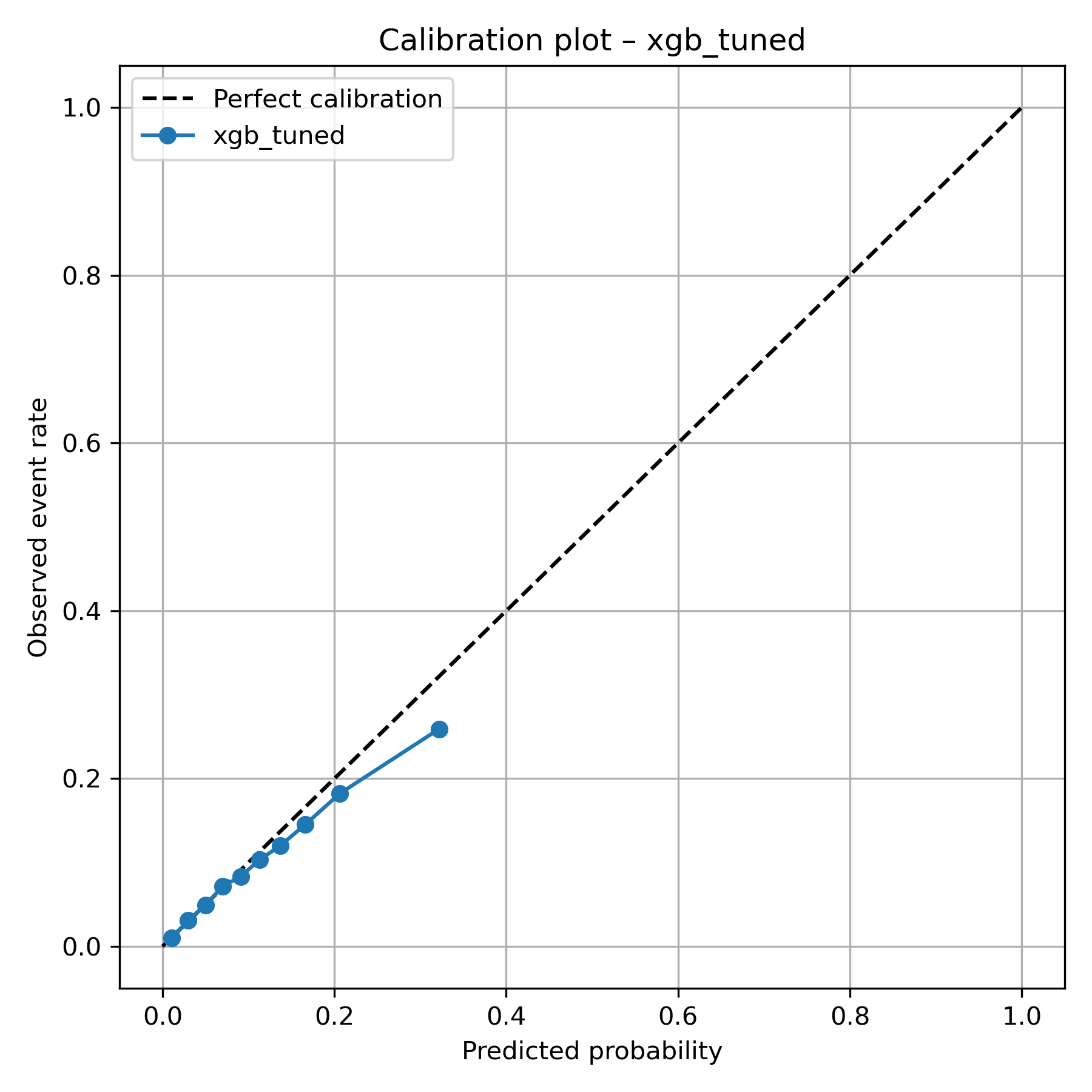

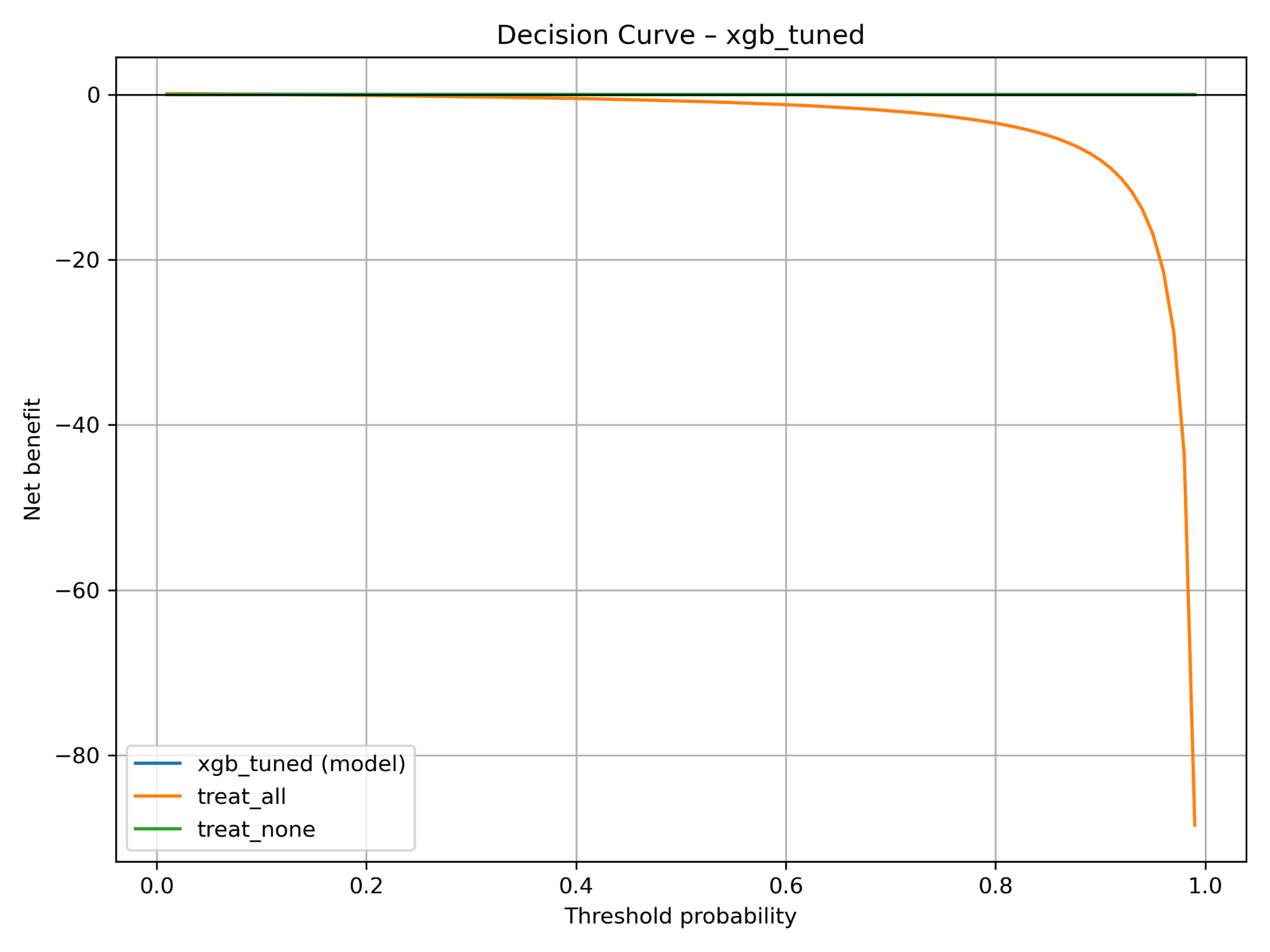

**( b )**

**( a )**

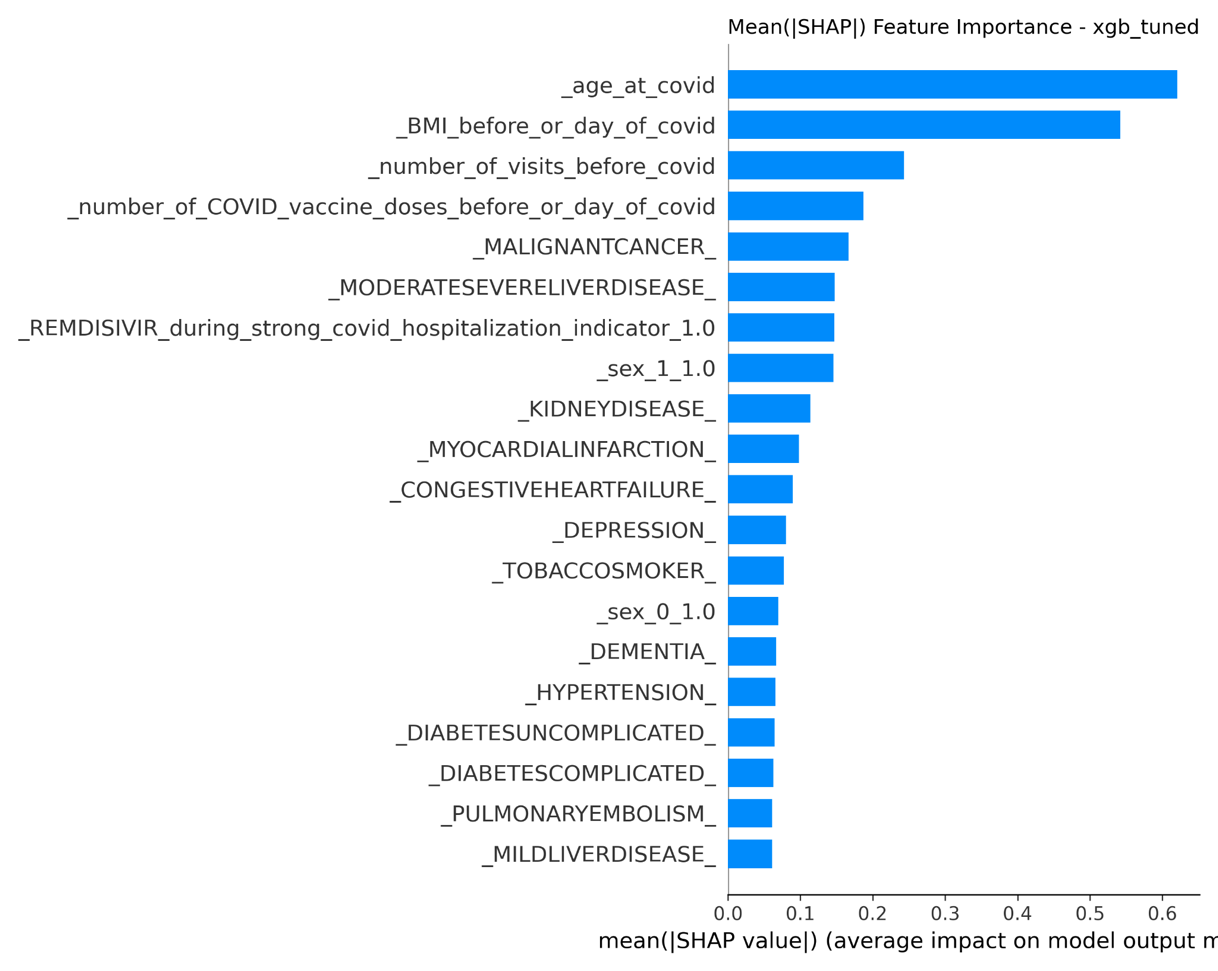

**( c )**

### S7 Fig. Multilayer perceptron (MLP) model (a) calibration plot, (b) decision curve, (c) training loss curve, and (d) mean SHAP feature importance for 60-day mortality with SMOTE (all patients).

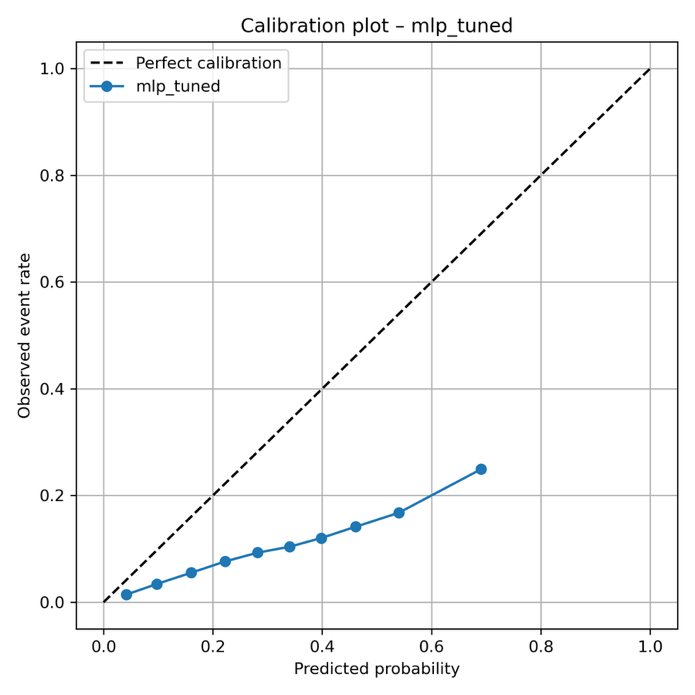

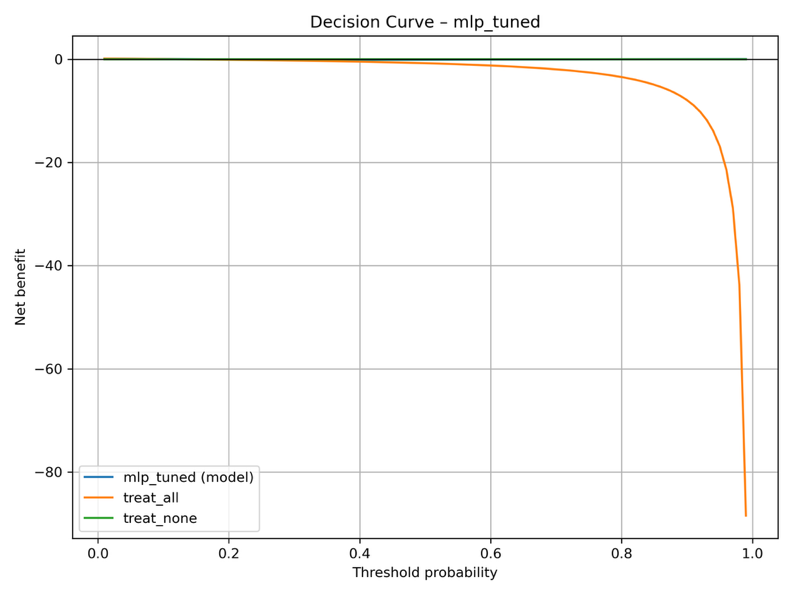

**( a )**

**( b )**

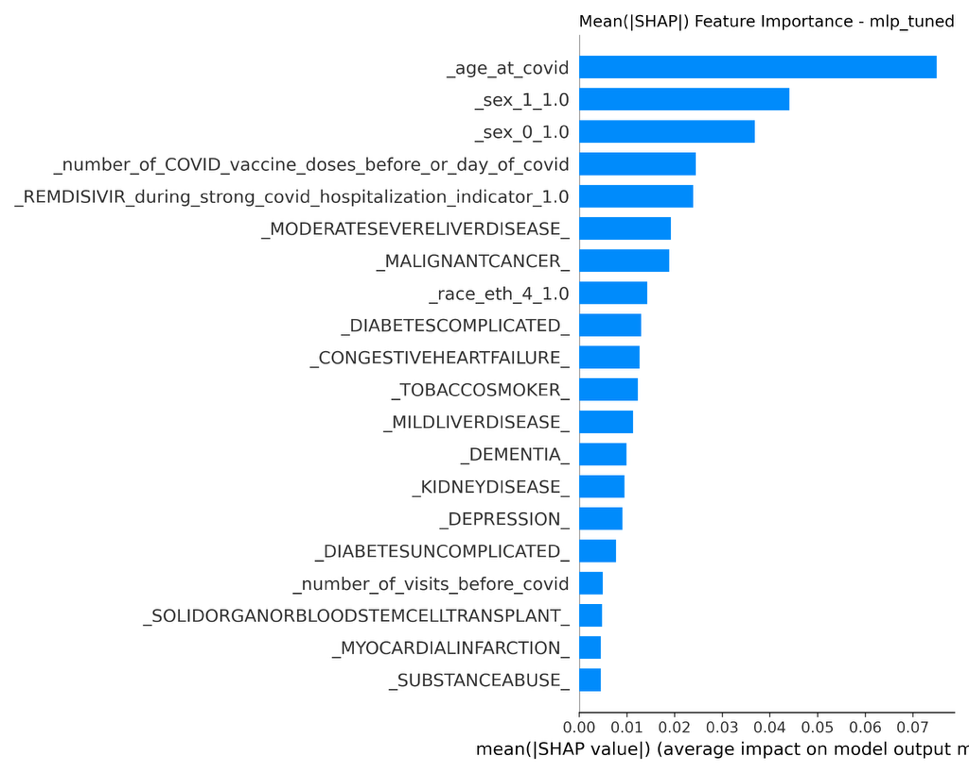

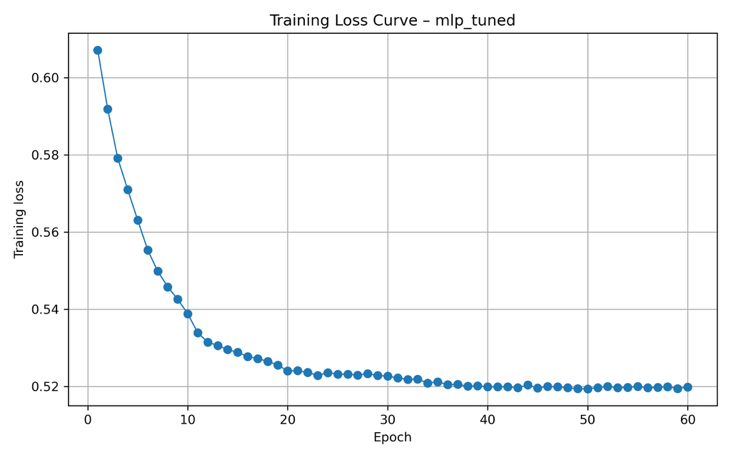

**( d )**

**( c )**

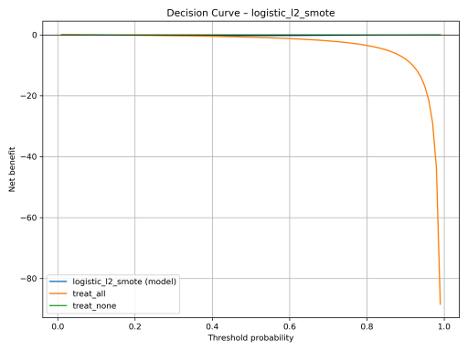

### S8 Fig. Logistic regression model (a) calibration plot, (b) decision curve, and (c) SHAP beeswarm plot for 60-day mortality with SMOTE (all patients).

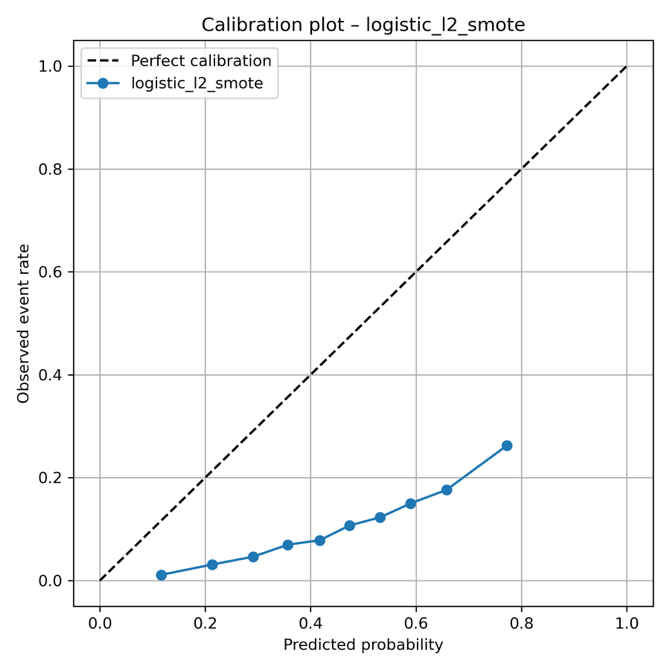

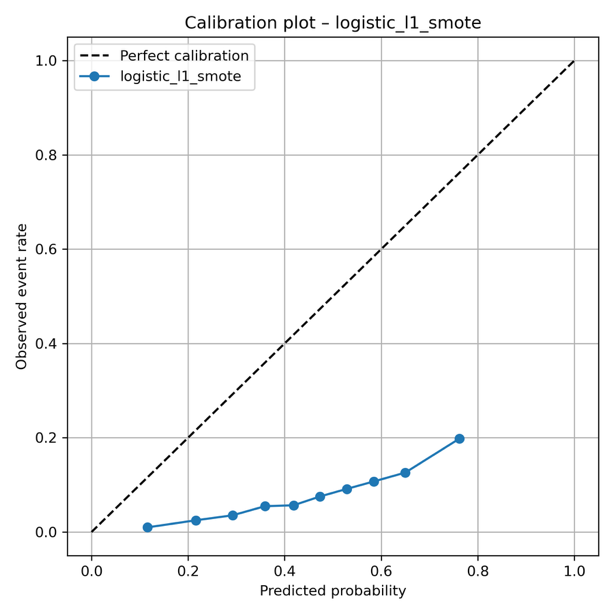

**( b )**

**( a )**

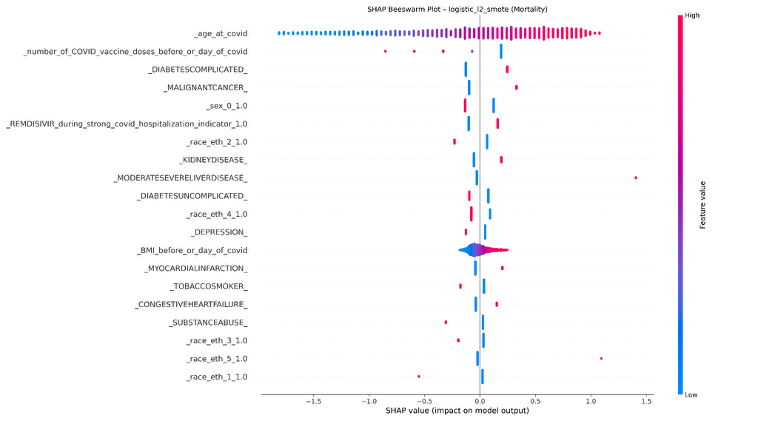

**( c )**

### S9 Fig. Elastic nets model (a) calibration plot, (b) diagnostic plot, and (c) mean SHAP feature importance for length of stay (all patients).

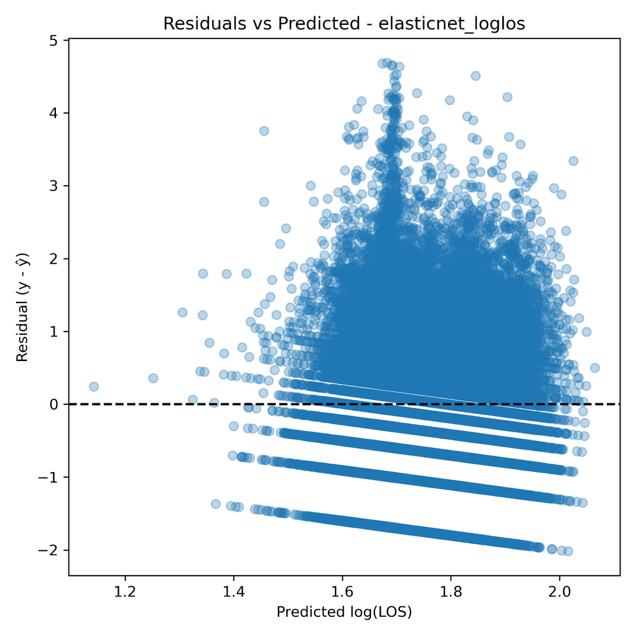

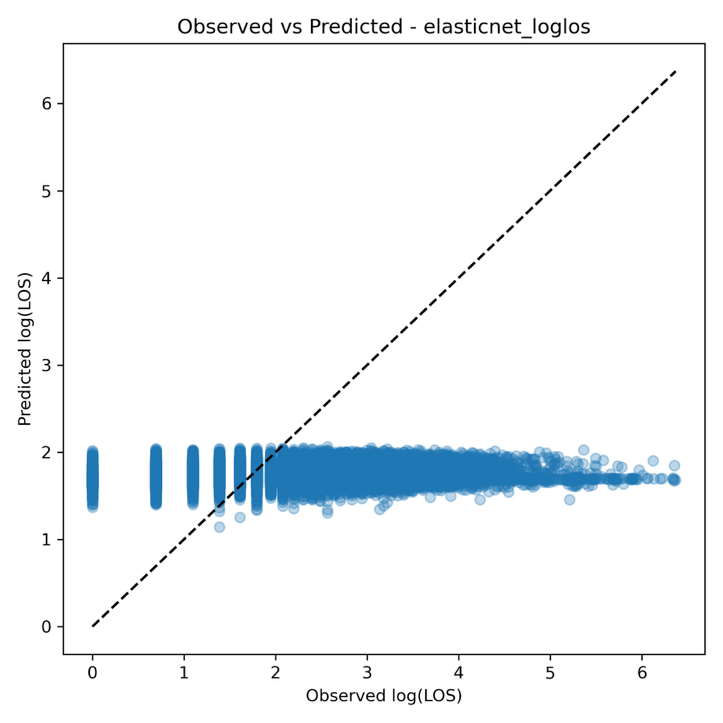

**( b )**

**( a )**

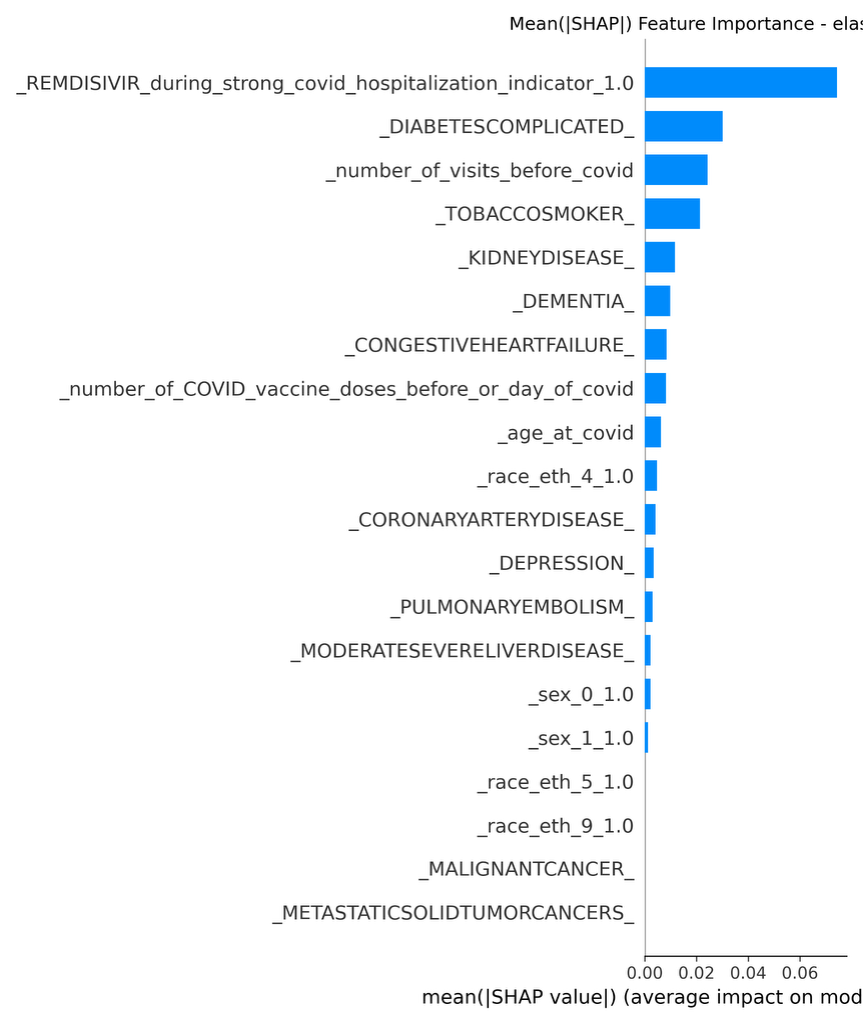

**( c )**

### S10 Fig. Random Forest model (a) calibration plot, (b) diagnostic plot, and (c) mean SHAP feature importance for length of stay (all patients).

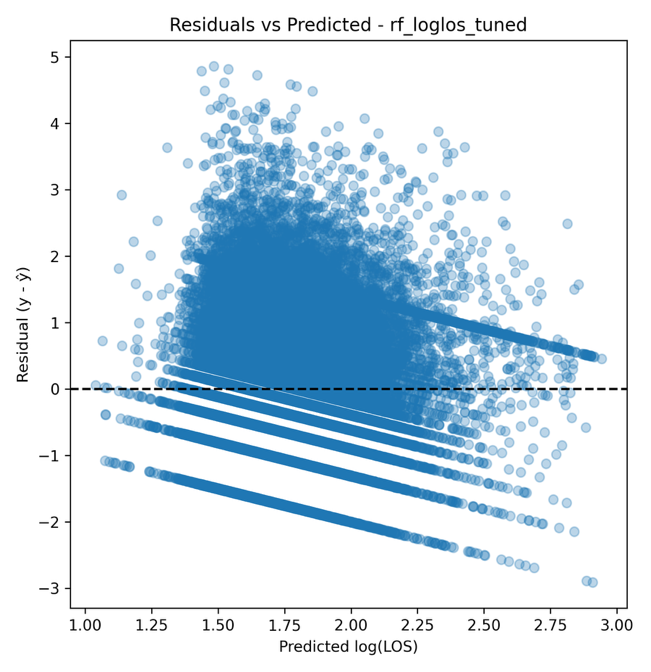

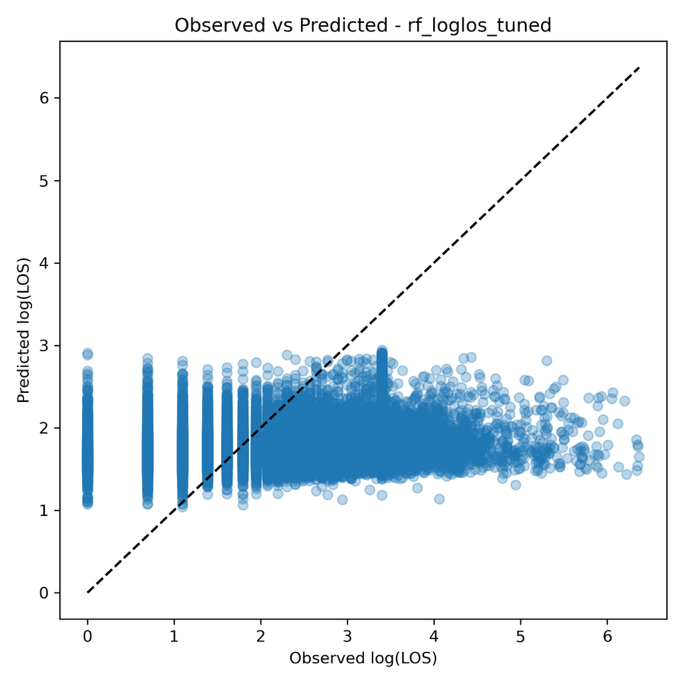

**( a )**

**( b )**

**( c )**

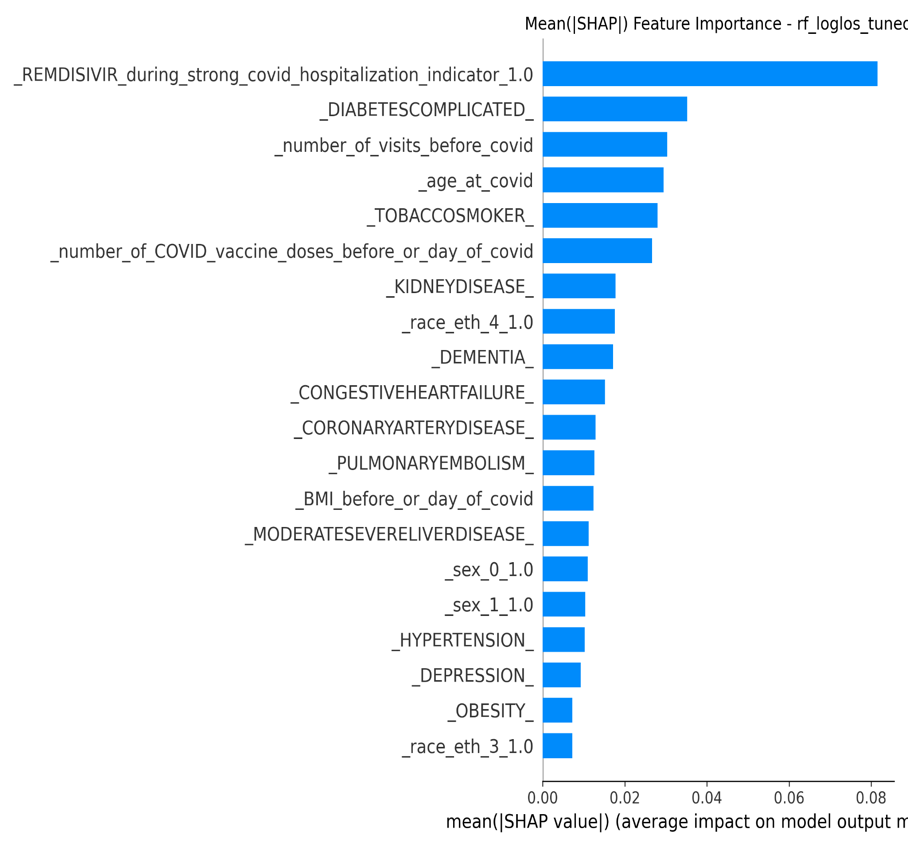

### S11 Fig. XGBoost model (a) calibration plot and (b) diagnostic plot for length of stay (all patients).

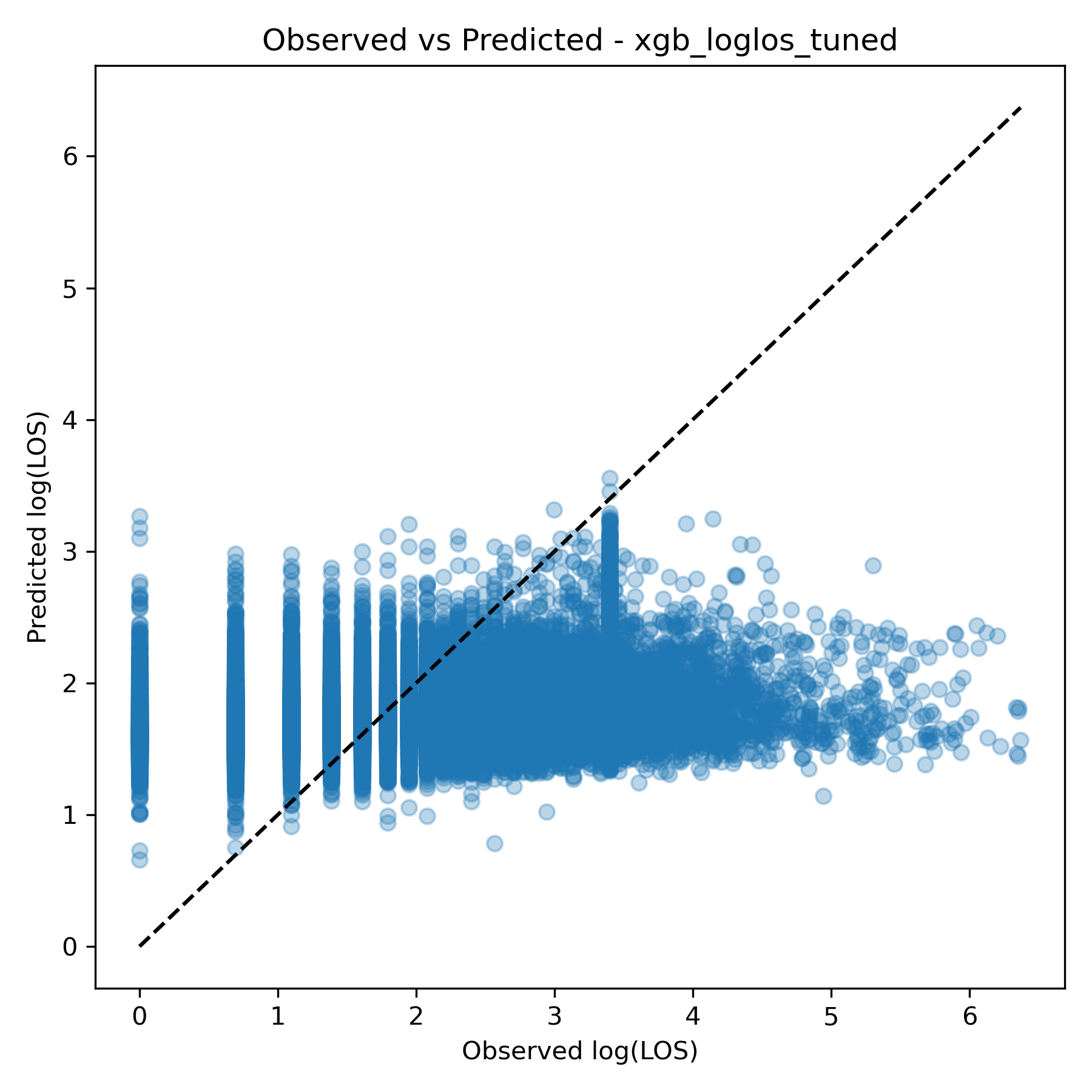

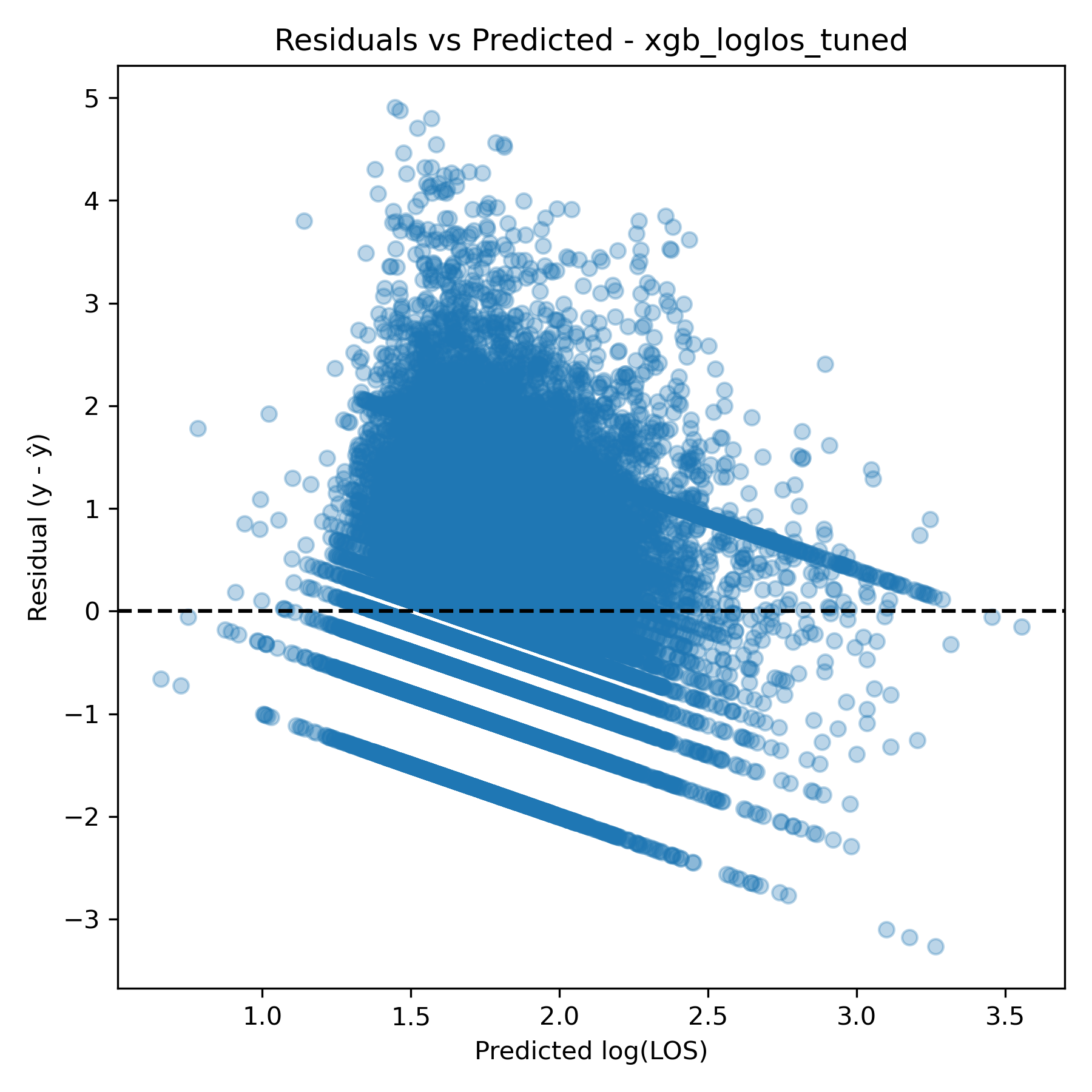

**( b )**

**( a )**

### S12 Fig. Multilayer perceptron (MLP) model plots on all patients for length of stay (all patients).

**Below are the following plots: (a) permutation feature importance for the MLP classifier, ranked by mean decrease in model accuracy on feature permutation, (b) partial dependence plots for number of COVID vaccines and number of visits before covid showing marginal effects on predicted outcome averaged across all observations, and (c) individual conditional expectation (ICE) plots for number of COVID vaccines and number of visits before covid displaying predicted outcome trajectories for individual observations as each feature varies.**

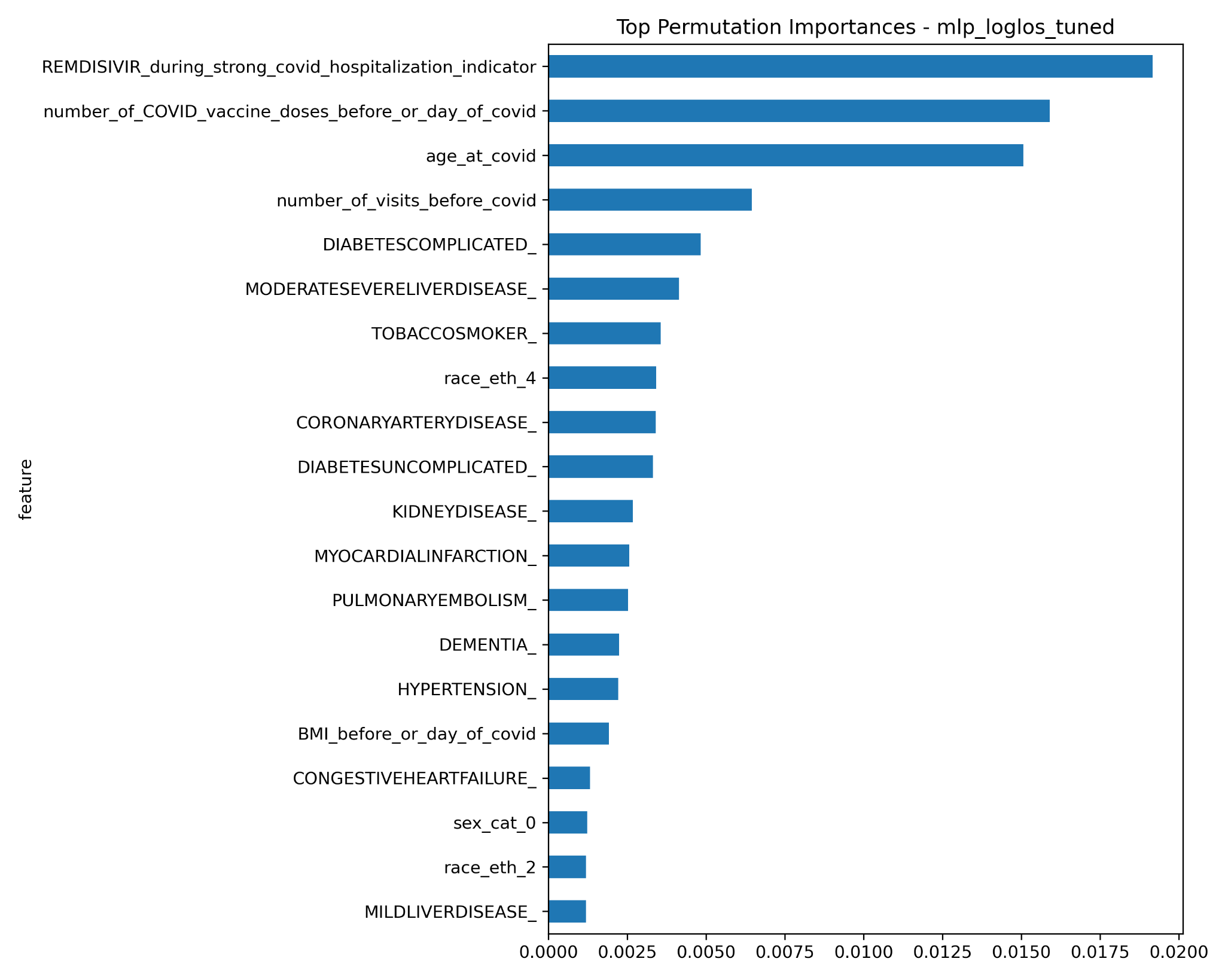

**( a )**

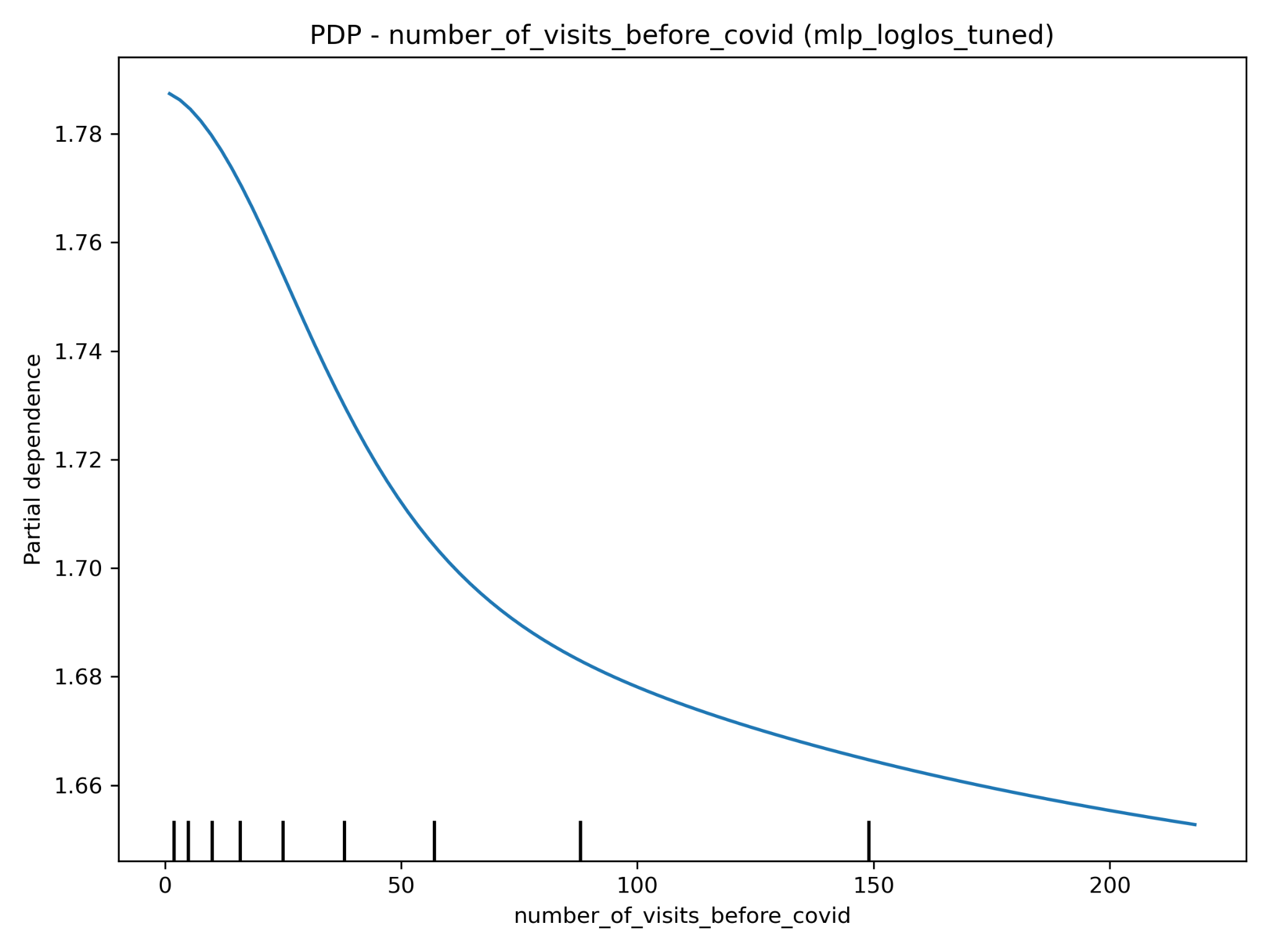

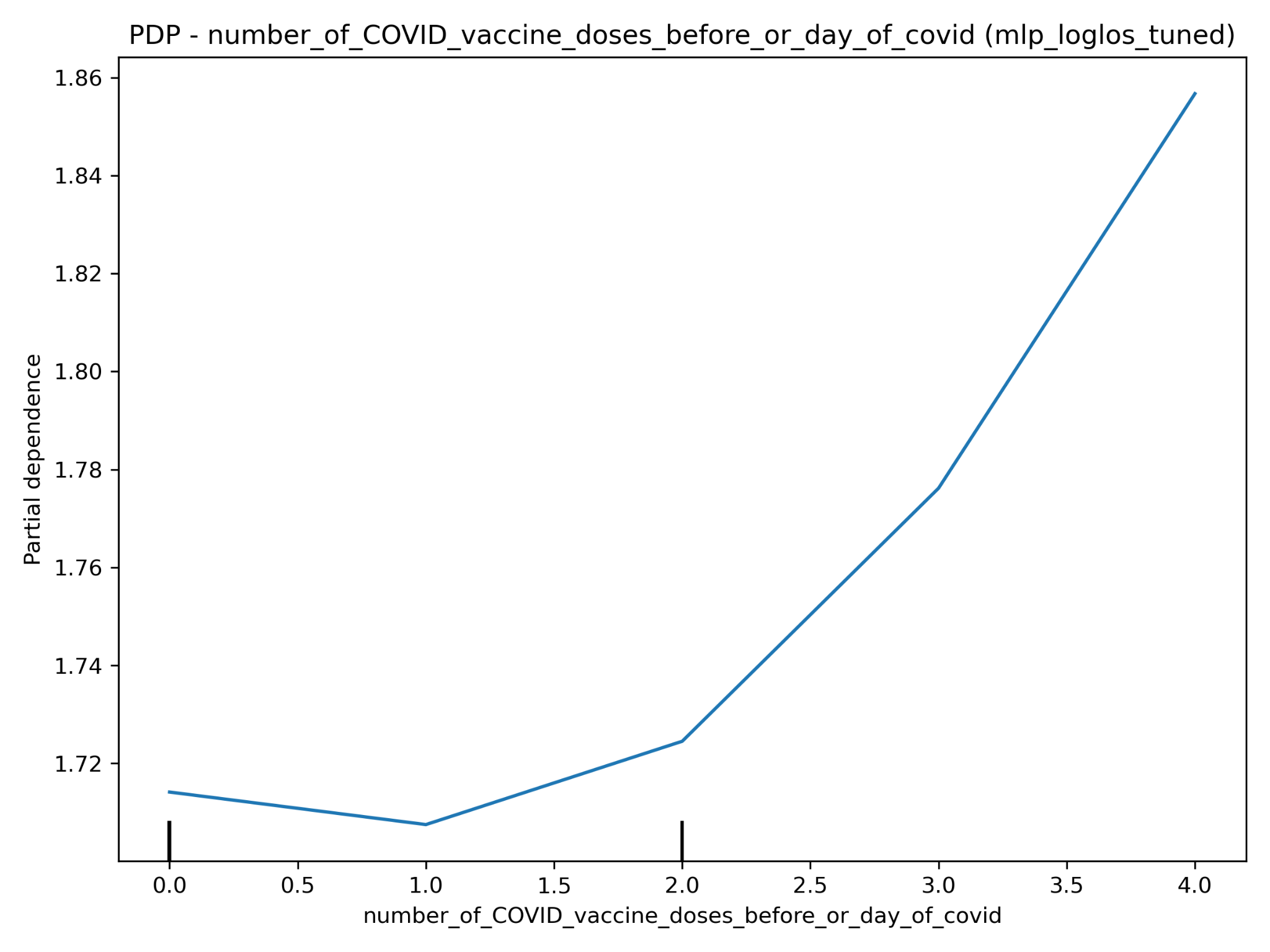

**( b )**

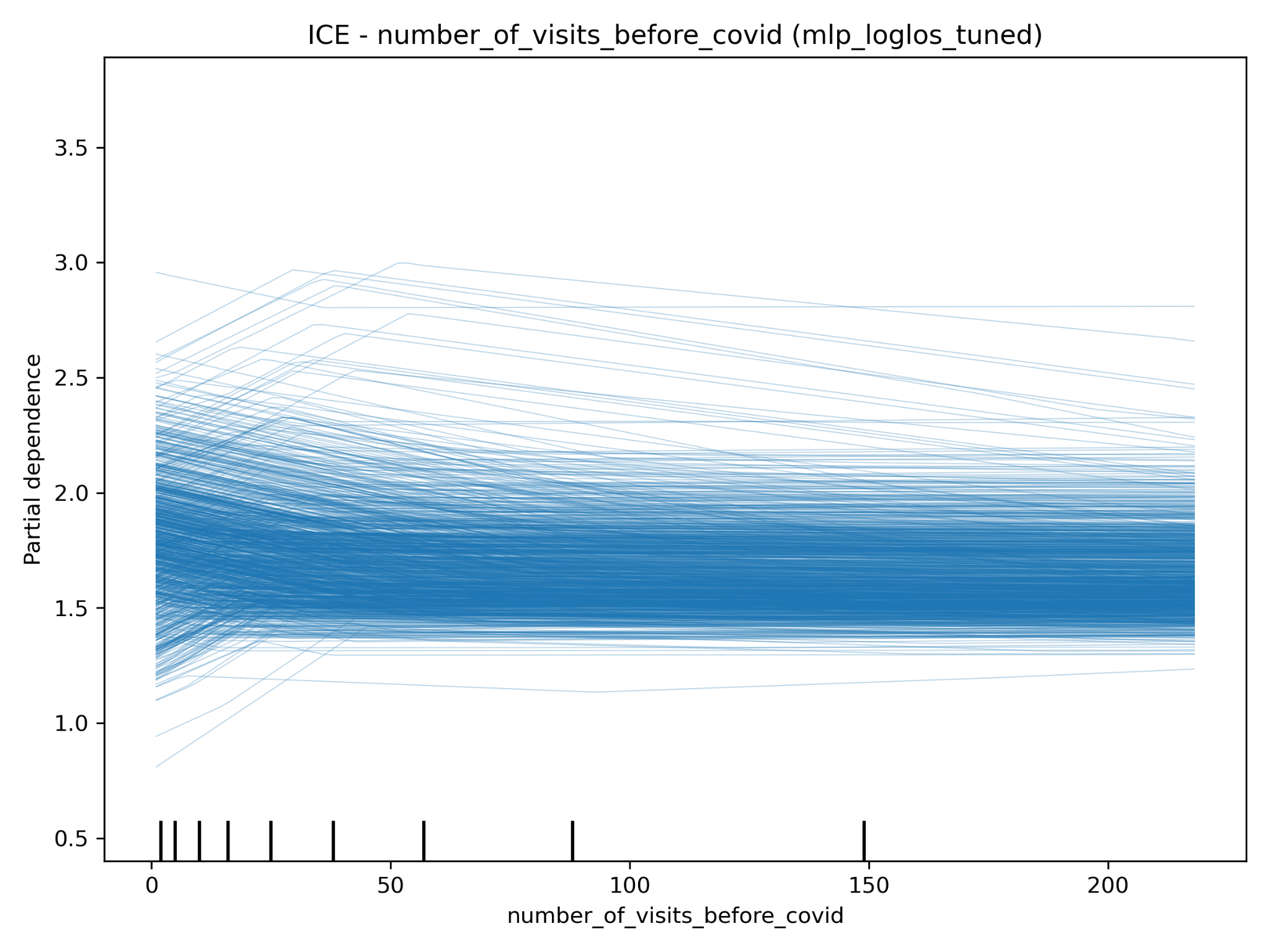

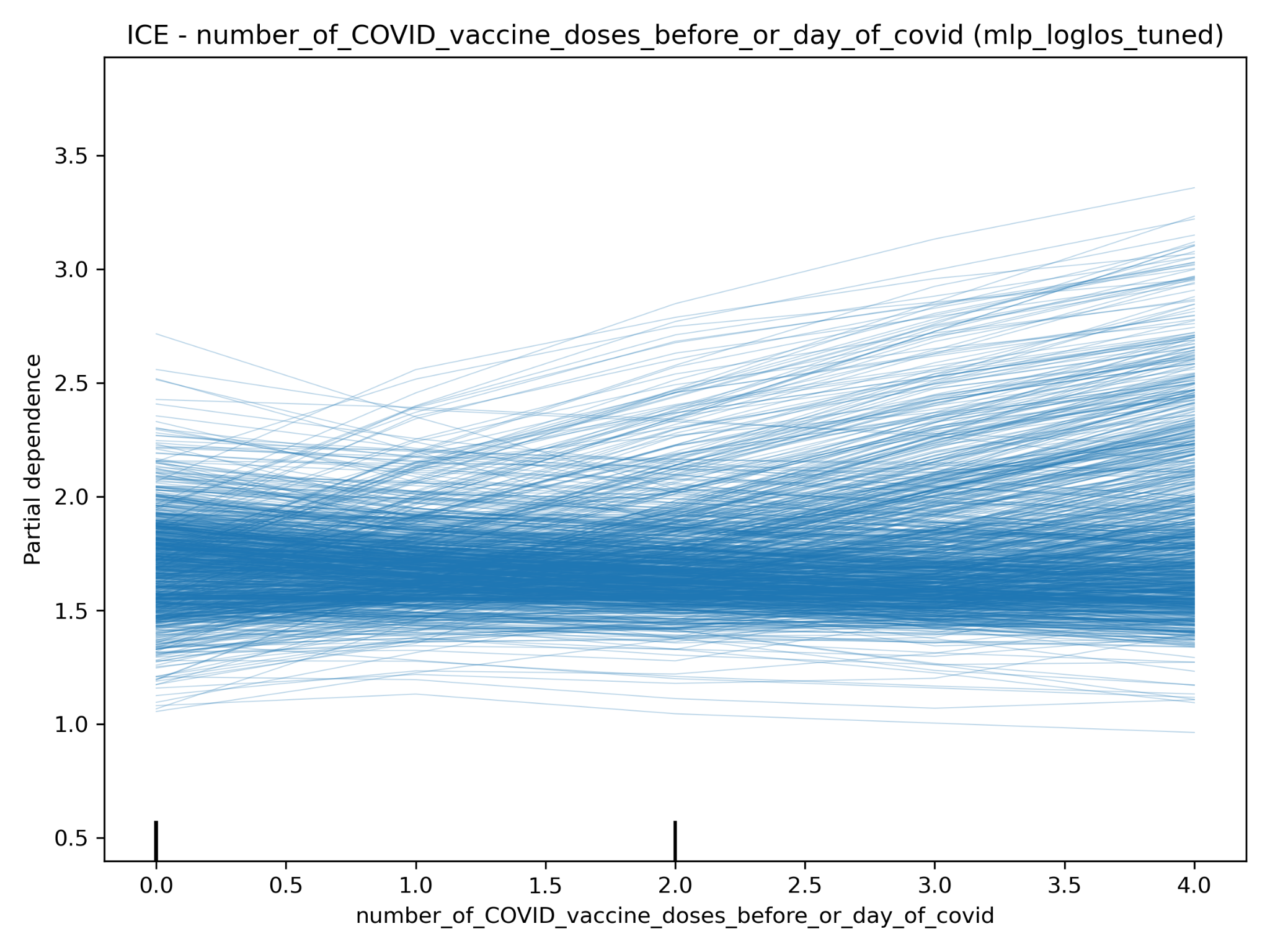

**( c )**

### S13 Fig. AUROC comparison of classification models for in-hospital mortality during COVID (all patients).

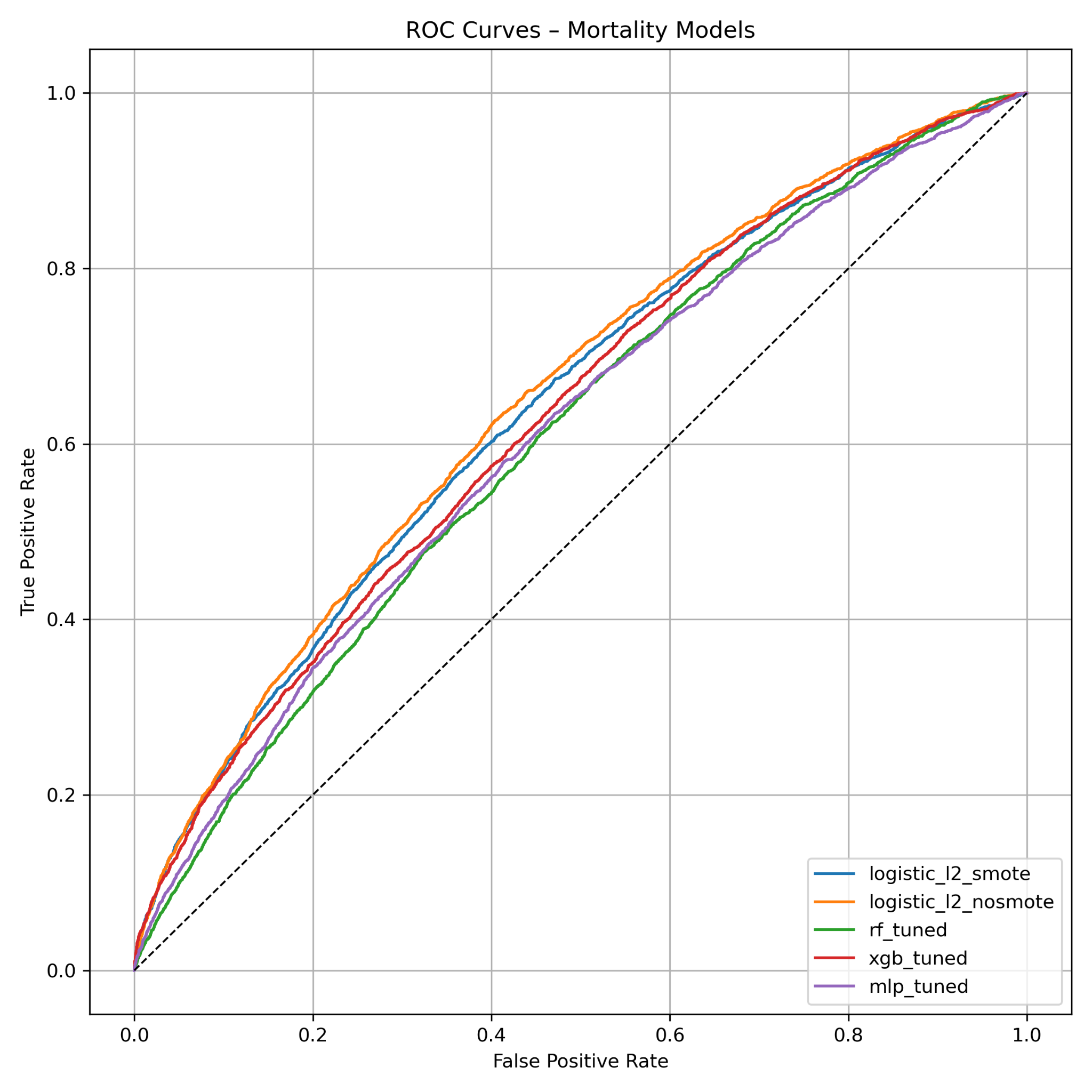

### S14 Fig. AUROC comparison of classification models for 60-day all-cause mortality on all patients.

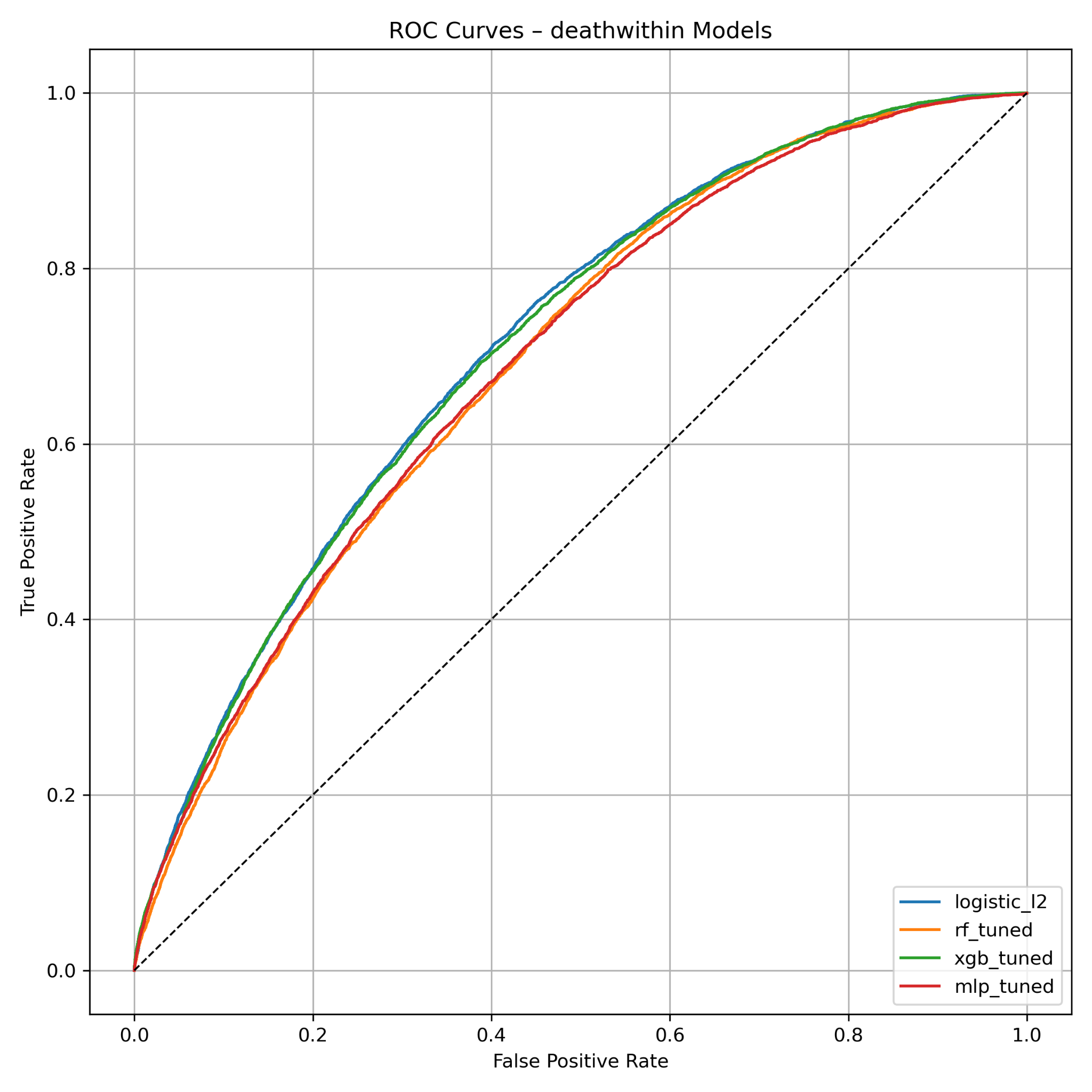
